# The topology of adolescent mental health

**DOI:** 10.64898/2026.07.13.26357465

**Authors:** Maria B. Jelen, Alexa Mousley, Kayson Fakhar, Estherina Trachtenberg, Yuankai He, Robert Kohler, Shambhavi Aggarwal, Varun Warrier, Danilo Bzdok, Sarah W. Yip, Duncan E. Astle

**Affiliations:** MRC Cognition and Brain Sciences Unit, University of Cambridge, Cambridge, UK; Institute of Computational Neuroscience, University Medical Center Hamburg-Eppendorf, Hamburg, Germany; Department of Psychology, University of Cambridge, Cambridge, UK; Department of Psychiatry, Yale University School of Medicine, New Haven, CT, USA; Department of Biomedical Engineering, The Neuro-Montreal Neurological Institute (MNI), McConnell Brain Imaging Centre (BIC), McGill University, Montreal, Quebec, Canada; Mila - Quebec Artificial Intelligence Institute, Montreal, Quebec, Canada; Department of Psychiatry, University of Cambridge, Cambridge, UK; Child Study Center, Yale University School of Medicine, New Haven, CT, USA; Center for Brain and Mind Health, Yale School of Medicine, New Haven, CT, USA; Wu Tsai Institute, Yale University, New Haven, CT, USA

## Abstract

The increased vulnerability to mental health problems in adolescence is frequently reported but poorly understood, hampered by a rigid diagnostic system which fails to capture intertwining symptoms and only loosely aligns with biological axes of variability. Here, we reconceptualised the mental health symptoms of young adolescents in the ABCD cohort (N=11862) as a latent topology of overlapping symptom dimensions, using an unsupervised machine learning algorithm to establish how transdiagnostic dimensions co-occur and overlap within individuals. Combining this with a novel classification approach, we delineated zones within this landscape, within which specific profiles of symptoms were robustly represented. These data-driven profiles were leveraged to establish associated resting-state functional connectivity and genetic characteristics. In doing so we recaptured the commonly reported p-factor axis as well as further symptom-subtype dimensions. Gene ontology analysis revealed that shared neurobiological and cellular mechanisms embedded in both the genome and transcriptome may confer risk for psychopathology.

## Main

Initiated by the onset of puberty, adolescence is a dynamic developmental period for the mind and brain, but also one of particular vulnerability for mental health difficulties, with most difficulties emerging in childhood or adolescence^1–3^. Multiple interacting developmental mechanisms contribute to this phenomenon, including plasticity in brain structure and function^4–6^, pubertal hormones^7^, genetics, social and cognitive changes^8^, regulation^9,10^ and adversity^11^.

One major challenge to reconciling these varied influences is accurately conceptualising and measuring psychopathology in the first place. Mental health presentations are frequently classified using phenomenologically derived, categorical diagnoses^12^. Despite the utility of this system, it is not best suited to understanding the developmental origins of psychopathology. Underlying biopsychosocial processes are not disorder-specific, with shared risk factors and biological correlates across multiple diagnoses^13^. Symptoms are often shared across conditions, and comorbidity is common^14,15^. Formal classification often misses or misclassifies individuals with atypical^16^ or ‘sub-threshold’ difficulties^17,18^. Developmental change provides further complexity: psychopathology changes across the lifespan, and individuals may arrive at similar phenotypes despite differing prerequisites (equifinality), or conversely share the same risk factors and diverge between pathology and resilience (multifinality)^19^. Taken together, the rigid categorical system artificially slices a continuum, masks dimensionality of symptomatology and dichotomises complex developmental change.

An alternative and increasingly popular transdiagnostic view posits that mental health problems can be understood as a set of continuous, biobehavioural dimensions^20^. Shared symptom domains are implicated across multiple diagnoses and are underpinned by shared mechanisms which do not fit neatly within traditional nosological divisions^21^. The treatment of psychopathology as more extreme presentations of features shared across the population also enriches analysis with a wider spectrum of real-life phenotypes and does not rely on strict boundaries of “health” versus “dysfunction”. This perspective has gained significant multidisciplinary attention including updated taxonomy (e.g. HiTOP^18^) and research practices (e.g. RDoC^22,23^).

A key aspiration of transdiagnostic approaches is to better connect complex symptom presentations with underlying mechanisms. This is not to say that this alignment is simple; the search for genetic and neural biomarkers has painted a complex picture of distributed and heterogenous correlates of broader psychopathology subtypes^24^. Most genetic and neurobiological predictors converge on shared risks across multiple neurodevelopmental and psychiatric diagnoses^25^, with few diagnosis-specific candidate markers^26^. Psychopathology dimensions have been associated with atypical structural^27,28^ and functional neural features^25,29–32^, often found in the longer-developing association cortex.^6^ One interpretation is that early neural markers of psychopathology reflect deviations from normative maturation of the brain^1,33,34^. Analogously, molecular genetics has suggested that psychopathology is highly polygenic and pleiotropic^35–38^, with cumulative risk emerging from multiple single-nucleotide polymorphisms (SNPs) of low effect size individually. Put simply, there is no one-to-one mapping between aetiology and singular diagnoses in the data, reflecting the multifactorial influence of biological, psychological and sociocultural factors^13^. Instead, biological mechanisms confer general, non-specific psychopathology risk, and some distinct patterning associated with continuous symptom domains.

While the transdiagnostic view is a promising perspective-shift for understanding psychopathology, we are still left with the challenge: how do we characterise heterogenous mental health difficulties? Prior work has derived transdiagnostic symptom dimensions (e.g. through factor-analysis^18,39^), but while valuable for capturing population-level variables, it does not tell us how these dimensions produce individual phenotypes. Alternatively, clustering participants may result in empirical data-driven groupings, but this approach echoes some limitations of traditional diagnostics, including low and somewhat arbitrary separation between profiles^40,41^.

We propose an alternative *topological approach* to understanding psychopathology. Unlike variable-centric models, this topological approach does not discard individual-level insight and instead harnesses multivariate within-person patterning techniques to understand how transdiagnostic dimensions of psychopathology interact and intersect within individuals. By reconceptualising psychopathology as a series of overlapping two-dimensional distributions, we may be able to capture the continuous *but non-linear* combination of symptom distributions, whilst also mapping the more idiosyncratic overlap of those non-linear distributions as they manifest as actual profiles of difficulty within individuals. To achieve this, we used an unsupervised multivariate machine learning model to map these overlapping topologies, and then classification, rather than clustering, to identify characteristic profiles within the topological landscape. We then tested whether and how this topological reframing of psychopathology maps on to functional brain connectivity and polygenic risk. Finally, we investigated whether patterns of gene expression in psychopathology-relevant brain areas are implicated in biological pathways shared with genes conferring risk for psychopathology.

## Results

### Overview of analysis workflow

We wanted a large dataset, with good coverage of multiple aspects of psychopathology, and large individual differences for us to map. For these reasons we used the ABCD cohort, which captures a population normative sample, with psychopathology levels ranging from very low (81.12% in the non-clinical range for all measures), to very high (8.61% in the clinical range on one or more scales). To capture multivariate psychopathology, we used 10 symptom subscales: the Child Behavior Checklist^42^; *Mania* from the General Behavior Inventory^43^ and *Disorders of Initiating and Maintaining Sleep* from the Sleep Disturbance Scale for Children^44^ (see Methods - *Behavioural Measures)*.

To generate the topological maps of symptoms we trained an unsupervised machine learning model called self-organising map (SOM)^45,46^. Over training, each participant is iteratively matched to the node in the map they are closest to in Euclidean distance (their Best Matching Unit (BMU), updating the weights of the node and its neighbours. Ultimately, each node represents the profile of participants for whom that node was the BMU, and adjacent nodes reflect more similar profiles than distant nodes (Methods – *Self-organising map)*. To make sense of this topology, we created a series of simulated ‘surrogate’ profiles, which can be thought of as potential candidate profiles the map may be representing. We then implemented a systemically validated and pruned SOM profile assignment using a supervised multilabel classifier, permutation testing to ensure non-random grouping of topology regions and matching individual participants to psychopathology profiles based on their BMU (Methods - *Psychopathology profiles*). We applied Partial Least Squares (PLS) correlation to investigate shared covariance between psychopathology profile map and resting-state functional connectivity (rsFC) as well as 17 polygenic risk scores (PGR) for common mental health diagnoses. As the final step, we used comparative pathway enrichment analysis to investigate if psychopathology risk genes are associated with similar biological processes as genes preferentially expressed in brain regions associated with psychopathology, using whole-brain transcriptomic data.

### Topologies of psychopathology

After data cleaning resulting in retention of 11862 participants, the SOM was trained using 10 features from 10 symptom subscales^42–44^. The details of training and parameter specifications are presented in the Methods - *Self-organising map*.

#### Psychopathology Profiles

Once the SOM is trained, adjacent nodes of the map retain continuous weights, representing a latent distribution of psychopathology phenotypes. To interpret SOM topology while taking advantage of the dimensionality of the map, we generated 8 ‘surrogate’ profiles as candidate psychopathology profiles the map may be representing. The reason why we did not just apply a clustering algorithm to the map is that the topology is necessarily and intentionally continuous, meaning that cluster boundaries will be arbitrary (Figure S1). Instead, we opted for a classification approach to identify zones of the map where we could classify specific profiles, relative to all other profiles. For the classification we generated 8 sets of 500 simulated individuals based on the covariance matrix of the original dataset, simulating increased symptoms in various theory-driven combinations (e.g. Internalising profile). We then trained a multilabel classifier (Random Forest) on the simulated data and classified the weights of each node on the SOM to assess which, if any, of the profiles best described areas of the map. To ensure profile mapping of the topology was not random, we permuted the weights of the map and repeated the classification to determine the minimum number of adjacent nodes which constitute a non-random zone. Six robust psychopathology profiles were retained (Figure 2b) (see Methods - *Psychopathology profiles*).

**Figure 1.**
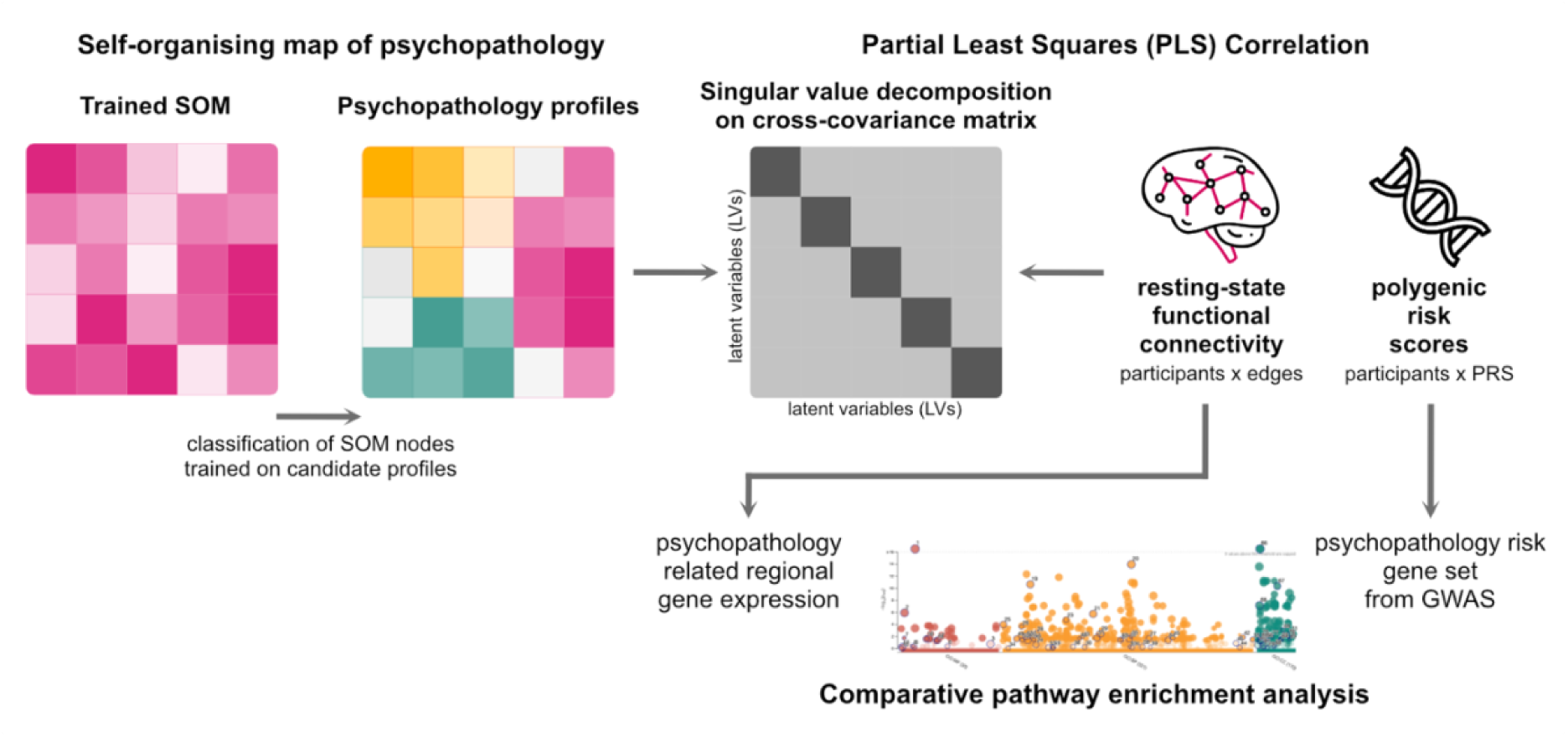
Schematic summary of analytic approach. The self-organising map (SOM) was trained on 10 symptom dimensions and partitioned using classification of nodes trained on candidate psychopathology profiles. The profile which best described a participant’s phenotype based on placement on the SOM was combined with i) resting state functional connectivity and ii) polygenic risk score in a partial least squares (PLS) correlation analysis, describing shared dimensions of covariance between biology and behaviour. The final step of the analysis involved comparative pathway enrichment analysis between psychopathology-related gene expression across the brain and previously established psychiatric risk genes from GWAS analyses.

**Figure 2.**
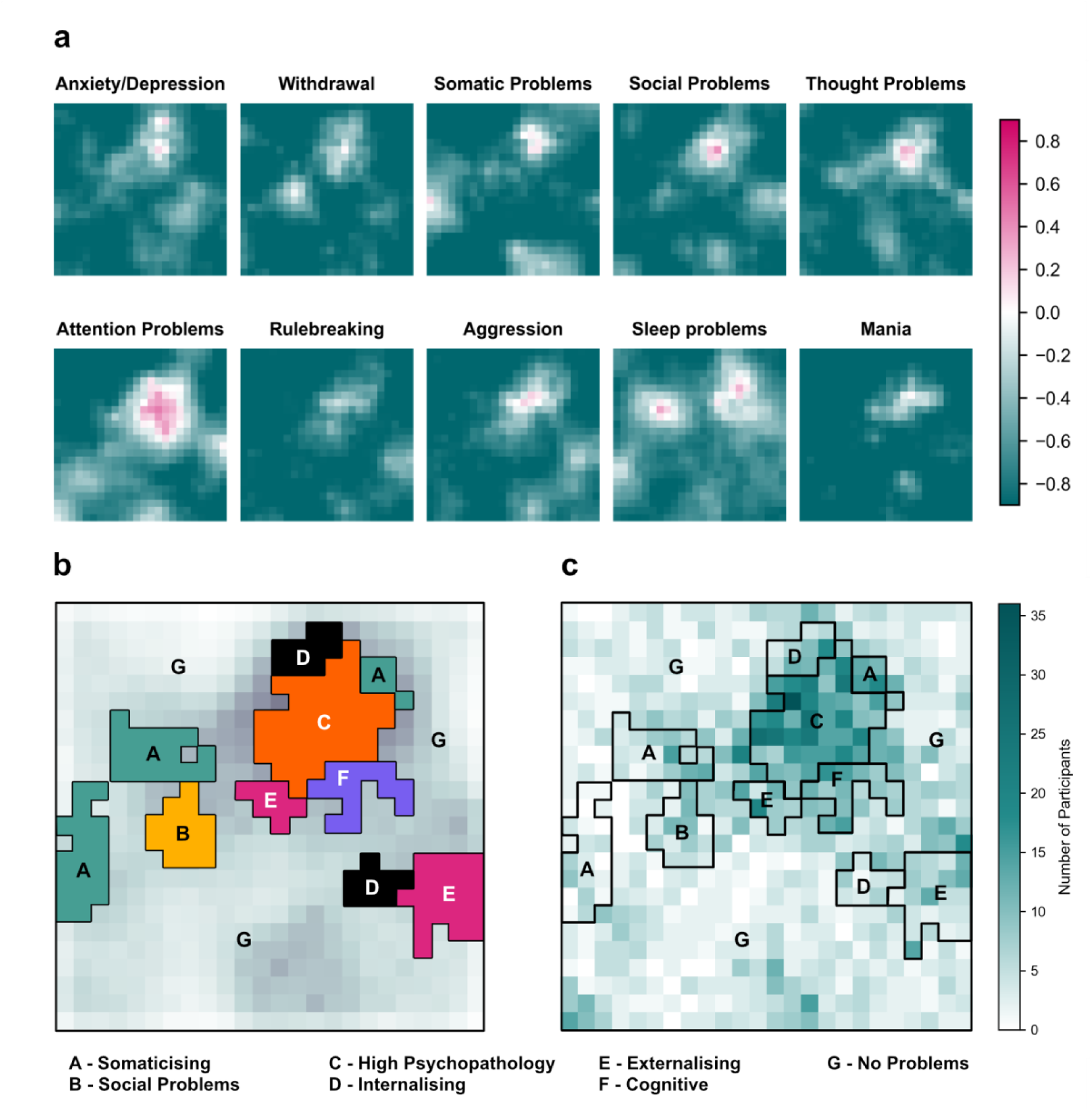
Topological map of psychopathology. **a.** Weight distribution from trained SOM, split by symptom scale. For each subscale, high weights (reflecting relatively higher symptom levels) are indicated in magenta and low weights (lower symptom level) are indicated in dark green. In the SOM projections, axes denote node position in network grid rather than a measure of independent variables. **b.** Intersections of psychopathology profiles on trained SOM. Profiles (A-G) which remained robust in validation and permutation testing are superimposed on top of trained SOM distances plot. **c.** Diagnosis density map imposed on SOM map grid. Data from all current diagnoses from parent-reported KSADS assessment were extracted for each participant and mapped back to SOM grid using BMU to display number of participants per node meeting the criteria for least one current KSADS-based diagnosis.

As validation step for the map, we sought to triangulate map topology with an alternative mental health measure. We therefore projected all current diagnoses from the Kiddie Schedule for Affective Disorders and Schizophrenia (KSADS^47^) scale (a semi-structured clinical diagnostic interview) onto the SOM grid; plotting the density of any current diagnoses for everyone mapped to an individual SOM node. The resulting map (Figure 2c) demonstrated similar topological peaks as symptom profiles, highlighting the relationship between heightened symptoms and likelihood of meeting KSADS diagnosis criteria.

We next tested how stable participants’ assignment to psychopathology profiles was across measurement time points, observing a high degree of stability conferred from high correlation between placement in the map and baseline and at follow-up time points (1-year, 2- year, 3-year and 4-year follow-up). Results are available in Supplementary Materials (Figure S2).

### PLS – Resting-state Functional Connectivity

In the first PLS model, we investigated the covariance between psychopathology profile membership (participants x binary profile membership) and resting-state functional connectivity (participants x edgewise connectivity) (see Methods **–** *Neurobiological modelling*). Importantly, PLS identifies patterns of covariance, rather than absolute connectivity differences, so it is important to note the sign of the loadings reflects association with the latent behavioural dimension and not increased/decreased functional connectivity directly. Significance of brain node loadings was established by masking the loadings with a bootstrap ratio (BSR) > 1.96, comparable to a significance threshold of p < 0.05^48^.

PLS revealed two significant latent variables (LV1: p >0.001, LV2: p = 0.012), explaining 28.3% and 18.6% of the covariance between rsFC and psychopathology profile membership. The first (LV1) expressed a pattern akin to the general psychopathology component or “p-factor”^49–51^, showing the No Symptoms profile as having a strong positive loading and the High Psychopathology, Externalising and Somaticising Problems profiles showing negative loading (listed in order of magnitude). In the brain pattern, positive loadings were associated with low symptom burden and involved strongest loading from connectivity within the default mode (DMN), salience/ventral attention and visual networks, as well as between dorsal attention (DAN) and visual networks (Figures 3, S3 in Supplementary Materials). The negative loadings corresponded to higher psychopathology and loaded primarily on the connectivity between the DMN and DAN, salience/ventral attention networks and visual, as well as between visual and somatosensory-motor networks.

**Figure 3.**
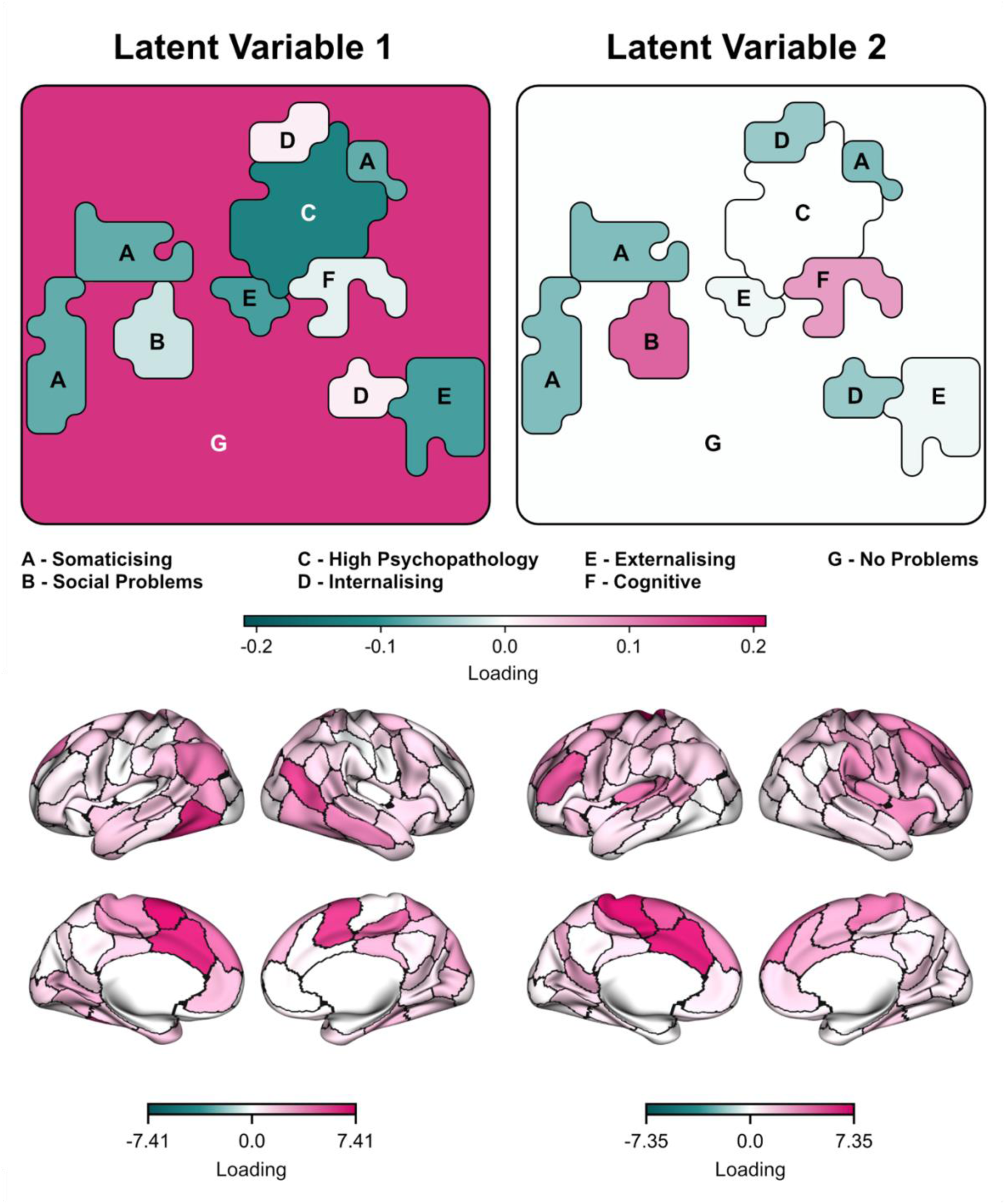
Significant Latent Variables from brain-psychopathology PLS. Upper panel shows loadings for LV1 (left) and LV2 (right) for psychopathology profiles derived from SOM, presented on approximation of SOM grid. LV1 pattern shows the positive loading of no psychopathology problems (G) and the negative loading of High Psychopathology (C) and other symptom domains. The orthogonal LV2 pattern examines the remaining covariance and represents an axis of symptom type. Lower panel shows signed nodal strength from PLS-derived loadings summed at node-level and projected onto brain surface, within LV1 (left) and LV2 (right), thresholded by bootstrap ratio and averaged across surface regions.

Orthogonally to LV1, the second latent variable (LV2) described a *symptom type axis*, with Social Problems and Cognitive profiles loading positively and Somaticising and Internalising profiles loading negatively. Individuals with higher social and cognitive problems had higher loadings on connections between the somatosensory-motor network and the default, control, and salience/ventral attention networks. People with greater levels of somaticising and internalising symptoms had higher loadings on connectivity between the DAN and visual, somatosensory-motor networks, and within-network connectivity in the DAN and DMN (Figures 3, S3).

To investigate the role of functional connections we identified in the PLS, we also investigated whether the stable (BSR-thresholded) edges implicated by the latent variables differ by *type* of connection between LV1 and LV2 (see Supplementary Materials – Results).

PLS results were robust in post-hoc analysis of potential confounding variables. A General Linear Model (GLM) predicting behavioural loadings from brain FC loadings and covariates including sex at birth, age, study site and fMRI motion, showed the FC loadings as significant predictors over covariate influence (Tables S2-3).

### PLS – Polygenic Risk Scores

In the analysis of covariance between polygenic scores and psychopathology profiles (N= 5672), we utilised 17 diagnosis PGRs: Attention-Deficit Hyperactivity Disorder (ADHD), Autism, Anorexia, Anxiety, Bipolar Disorder, Major Depressive Disorder, Obsessive-Compulsive Disorder (OCD), Panic Disorder, Post-traumatic Stress Disorder (PTSD), Schizophrenia and Substance Use Disorder (SUD). ADHD and Autism PGRs were further divided into diagnosis during childhood vs. adulthood; Bipolar score was divided into Type I and II. For this portion of genotyping analysis, the sample was constrained to participants with European ancestry.

Here PLS described three significant latent dimensions of covariance between the PGRs and psychopathology profiles, explaining (LV1: p >0.001, LV2: p = 0.005, LV3: 0.034), explaining 77.8%, 9.8% and 6.4% of covariance respectively.

LV1 delineated a “p-factor” axis in the behavioural variable, with symptom items including High Psychopathology, Externalising, Cognitive and Somaticising loading positively and No Problems loading negatively. On the genetic risk level, higher symptoms (p-factor loadings) were associated with higher risk scores across 11 PGRs, with Depression, PTSD, ADHD, Anxiety and Autism showing highest loading (Figure 4).

**Figure 4.**
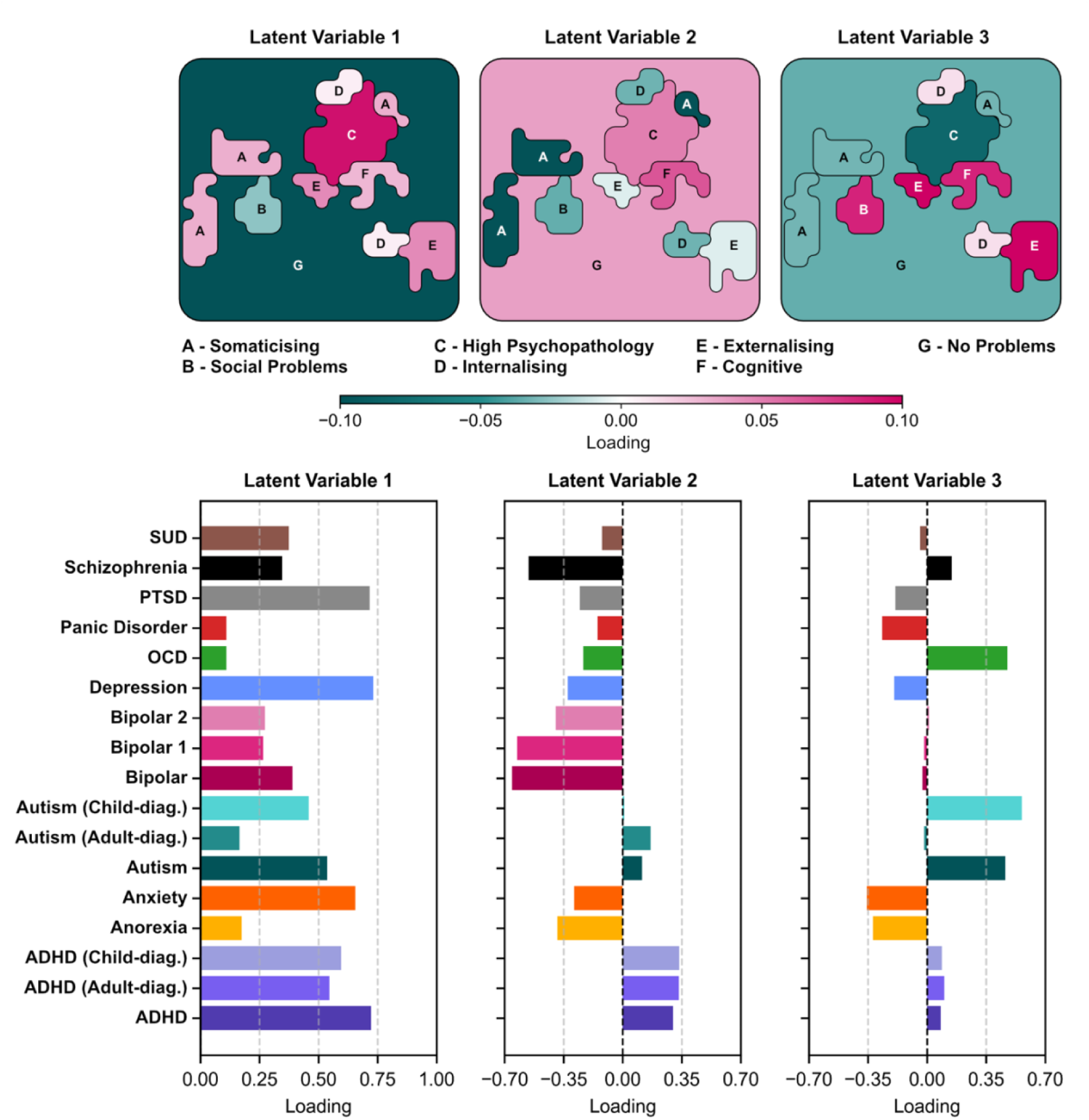
Significant Latent Variables from PGR-psychopathology PLS. Each vertical panel presents the magnitude of PLS-derived loading on each feature within the three significant latent variables (LV1-3). The top plots represent behavioural loadings while the bottom plots present loadings on PGRs.

LV2 contrasted more classic psychopathology (Somaticising, Internalising, Social Problems subscales) with a cognitive symptom axis, expressed in positive loadings primarily from the Cognitive profile. In combination with PGR loadings, the positive loading of the LV may be interpreted as neurodivergence, demonstrated with positive loadings of ADHD and Autism polygenic scores. Conversely, PGRs for more classically construed psychopathology, including Bipolar, Schizophrenia, Anorexia, Anxiety and Depression, loaded negatively, aligned with behavioural weightings.

LV3 showed a more distributed pattern of loadings that may be interpreted as an expression of broader internalising difficulties contrasted with more specific, cognition-related symptoms. Lower loadings were associated with increased scores on High Psychopathology, covariant in the PGRs with Anxiety, Anorexia, Panic Disorder, PTSD and Depression risk. Higher symptoms on Externalising, Cognitive and Social Problems subscales reflected higher loadings on Autism, OCD and Schizophrenia.

The genomic PLS results were robust to potential confounding variables assessed using GLM (Supplementary Tables S4-5) and the effects of population stratification, assessed by repeating the PLS with residualised PGRs after regressing out 20 genetic principal components (Figure S6).

We additionally observed that the latent variable explaining most covariance (LV1) in both gene-behaviour and brain-behaviour PLS appeared to recapitulate the principal p-factor^49^. The correlation between the participant-level behavioural scores for LV1 for the gene and brain connectivity PLS analyses was high and significant (*ρ*(2160) = −0.994, p < 0.001). Note, the direction of PLS loading assignment is consistent within a latent variable but positive/negative assignment is arbitrary. Put simply, the first LV1 that links behaviour with either (i) functional brain connectivity, and (ii) genetic liability, is virtually identical. There was also a low positive correlation between the second LV in gene-behaviour and fMRI-behaviour PLS (*ρ(*2160) = 0.162, p < 0.001).

### Overlapping gene ontology between psychopathology-relevant transcriptome and genome

What do the genes in the polygenic scores do? And why are they linked to a similar set of psychopathology topologies as a distributed pattern of resting-state functional connectivity? One possibility is that there is a set of shared biological processes that have a regional expression profile in the brain. We tested this possibility in a final analysis.

We first obtained processed regional gene expression patterns from the Allen Human Brain Atlas (AHBA; https://human.brain-map.org/), containing expression of 13561 genes mapped to 50 left hemisphere nodes in the matched Shaefer-100 atlas. We then computed regional differences in functional connectivity based on the contrast identified by our rsFC PLS, taking the mean difference between connectomes of individuals in the “No Problems” profile and those in “High Psychopathology”, “Externalising” and “Somaticising” profiles. This resulted in a vector of psychopathology-related connectivity contrasts across 100 nodes, from which we retained 50 left-hemisphere nodes to match AHBA. These two matrices were the input for a PLS analysis, which identified one significant latent dimension (p = 0.04) of shared covariance. Based on a BSR threshold of 2.58 (p < 0.01), we extracted 1634 stable genes loading on the latent variable.

Additionally, we extracted the PGRs significantly loading on the first psychopathology LV (ADHD, Anxiety, Autism, Bipolar, Bipolar Type 2, MDD, PTSD, SUD) and consulted the Genome Wide Association Studies (GWAS) based on which the PGRs were calculated to identify significantly implicated genes (see Supplementary Table S7). After removing overlap, 570 unique genes were identified as relevant for risk for mental health diagnoses.

We then used multi-query comparative pathway enrichment analysis implemented in g:Profiler to test if 1) genes reflecting psychopathology-relevant transcription patterns and 2) genes associated with formal diagnoses from PGR/GWAS converge on shared biological ontologies including biological processes (BP), molecular functions (MF) and cellular components (CC) (see Methods – *Gene Expression and Ontology*).

Enriched genes identified in transcriptional and risk gene sets were not random but instead converged on 180 gene ontologies significantly enriched in both gene sets (*p_adj_* < .05), primarily including biological processes (67.22%). Across both gene sets, the most prominent shared enrichment for BP included system and cellular organisation (32 BPs, expression *p_adj_* < 0.049, GWAS *p_adj_* < 0.031) and neurogenesis (27 BPs, expression *p_adj_* < 0.043, GWAS *p_adj_* < 1.88 x 10^−5^) (Figure 5a). For MF, protein binding (6 MFs, expression *p_adj_* < 0.032, GWAS *p_adj_* < 6.73 x 10^−3^) emerged as most significantly enriched in both lists (Figure 5b). In terms of CC, shared enrichment was highest for cell organelle (16 CCs, expression *p_adj_* < 0.034, GWAS *p_adj_* < 0.049), cell body projection (14 CCs, expression *p_adj_* < 0.041, GWAS *p_adj_* < 0.005) and synapse junction (13 CCs, expression *p_adj_* < 0.034, GWAS *p_adj_* < 0.012) (Figure 5c).

**Figure 5.**
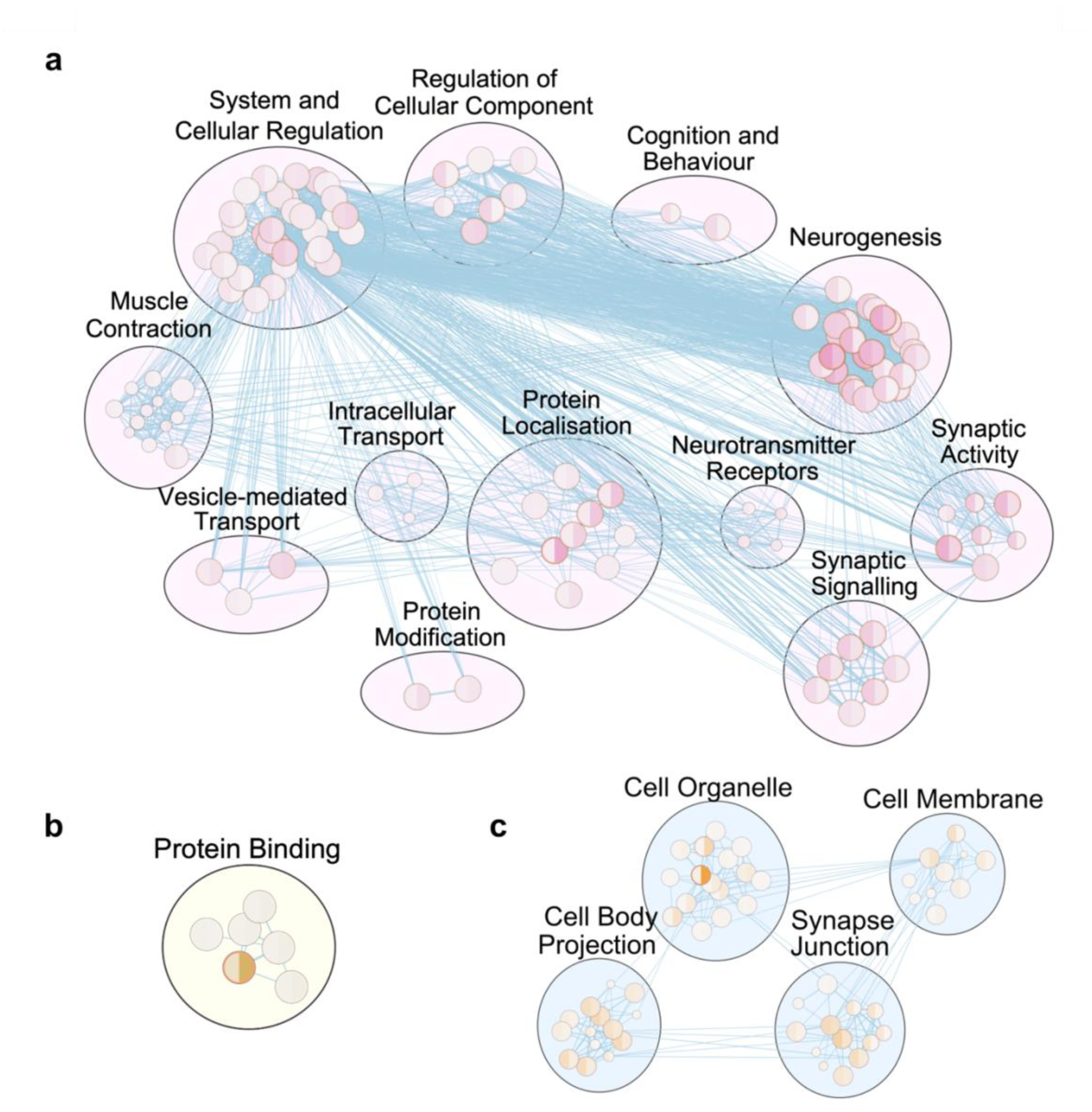
Psychopathology-relevant regional gene expression and genetic risk for mental health diagnoses converge on shared biological processes, molecular functions and cellular processes. Clusters delineate annotated gene function groups. Nodes represent individual biological processes identified as enriched in GO analysis, with size reflecting number of contributing genes and shade representing enrichment in respective gene lists (left side of sphere - GWAS, right side of sphere - expression; darker shade = higher enrichment). Blue edges represent degree of overlap between biological pathways based on similarity coefficient. **a.** Biological processes. **b.** Molecular functions. **c.** Cellular components.

## Discussion

We used a topological framework to reconceptualise distributions and profiles of mental health in the ABCD Study^®^. Unsupervised self-organising maps represented a dimensional landscape of psychopathology from multivariate symptom combinations, stepping beyond diagnostic categories to instead capture dimensional intersections within young individuals.

We identified interpretable symptom profiles within this landscape and investigated the covariance between young adolescents’ psychopathology and biology. Comparative gene enrichment analysis suggested the risk for psychopathology is underpinned by biological mechanisms embedded in both the genome and transcriptome.

Firstly, we were able to observe a general division in the SOM landscape relating to the differentiation between low symptoms and any kind of elevated psychopathology, observable in topological areas where weights of multiple symptom features overlapped (Figure 2a). Secondly, we found that our map can capture previously reported dimensions in child/adolescent mental health, including internalising and externalising difficulties (e.g. spectra within HiTOP^18^, CFA-derived factors^52^, item and exploratory factor analysis^53^), alongside previously described transdiagnostic factors, including Cognitive and Somatic dimensions^52–54^.

Beyond recapturing the previously described p-factor and psychopathology dimensions, our novel approach provided allowed individuals to be mapped back to places within SOM topology. This allowed us to draw interpretable links between multivariate psychopathology profiling and individual neurobiology and genetics without reduction to group-average factors or constructs. As opposed to clustering, our approach does not impose a separation of what we understand to be overlapping and interacting symptom dimensions. Instead, the latent topology allows for the investigation of a many-to-many mapping between symptoms and biological features^55^.

Additionally, we have moved beyond a case-control dichotomy and utilised the full spectrum of mental health difficulties, including moderate or subthreshold difficulties. Measuring participants at a very young age (preceding or entering puberty) allowed us to observe a developmental period when many problematic symptoms are only just emerging. The non-linearity of the SOM also meant the map was robust to “atypical” combinations of symptoms, without requiring clear linear interdependencies. Complex mental health presentations often fall out of traditional diagnostic classifications and associated clinical pathways^16^. In contrast, assessing where a young person falls in a complex symptom space has potential to contextualise the unique phenotype and retain individual links to biological and environmental covariates.

The first latent dimension in rsFC and polygenic risk score PLS was highly correlated and reiterated the frequently reported p-factor of psychopathology^49–51,56–58^ differentiating individuals along a global psychopathology gradient and suggesting the p-factor is a higher-order organising factor within both genetic and neurobiological covariates of mental health.

Further latent variables informed the next level in the hierarchy based on symptom type. In our rsFC PLS analysis, connections covarying with higher psychopathology included higher-order networks including the DMN, DAN and salience/ventral attention, previously associated with psychopathology^29,59,60^. Higher-order executive networks were also associated with symptom dimensions in LV2, with increased involvement the anterior and dorsal prefrontal cortex, regions critical for cognitive control and goal-directed behaviour^61^. The negative loading, associated with internalising and somaticising symptoms, was differentiated by a higher impact of connectivity within the DAN and DMN, both implicated in depression^62–64^. Although many of the same networks emerged as contributing significantly to both LV1 and LV2, PLS ensures the latent factors are orthogonal, indicating that these networks are important for more specific symptom-type connectivity patterns after accounting for p-factor covariance. Investigating the directionality of connectivity and multimodal modelling involving other features of brain organisation will be important future steps.

In the PGR PLS, subsequent latent variables (LV2 and LV3) mapped onto symptom-type subdivisions. However, rather than a one-to-one mapping, groups of PGRs contributed to axes of symptoms, highlighting the polygenic nature of psychopathology. For example, LV2 differentiated mood and behavioural psychopathology from neurodevelopmental difficulties, visible in positively loading PGRs (Autism, ADHD) associated with Cognitive symptoms. LV3 explained less of the covariance in the model but the combination of PGRs related to Autism, OCD and Schizophrenia could speculatively constitute an extended neurodevelopmental axis on which schizophrenia is sometimes placed^65,66^. The covarying psychopathology profiles of Cognitive and Externalising problems could relate to shared domains such as inflexible thought patterns or repetitive behaviours^67^. Considering the high degree of risk overlap, future research to disentangle general versus specific polygenic risk factors is worthwhile. At present, our analysis seems to support a hierarchical risk model, with general p-factor risk and more specific transdiagnostic symptom-type ordering^36^.

The implications of the covariance between dimensional symptoms and diagnosis-based PGR are also worth considering. PGR scores are derived from GWAS findings, which stem from a binary diagnosis mapping (case-control diagnostic division) in adult samples. Here, they meaningfully covary with transdiagnostic symptom profiles in a vastly different sample of normative young adolescents. This suggests that rather than carrying specific risk for a psychiatric diagnosis, polygenic risk factors may instead influence variability in common biological pathways which, in some combinations, result in heightened psychopathology risk.

Finally, we observed overlapping biological, cellular and molecular pathways between psychopathology-related regional transcription and mental health risk genes. Many of these processes relate to brain development and maturation (e.g. neurogenesis), brain functioning (e.g. neurotransmitter system, synaptic signalling) and system regulation (e.g. membrane transport and protein localisation). Many of these processes, such as altered neurotransmission and synaptic organisation, have been observed in multiple types of psychopathology^68–71^. Atypical neurodevelopmental processes captured at molecular, connectomic and systems levels are a major hypothesis for the aetiology of mental health difficulties^1,33,34^. However, generally the pathways we have observed are in themselves normative and necessary mechanisms for brain development and functioning. This supports the view of psychopathology not as an aberrant and binary occurrence, but instead a combination of deviations from typical maturation or functioning of the brain. This in turn suggests that they are part of normative developmental mechanisms which may malfunction when risk factors accumulate, differentiating the level and type of psychopathology the young individual is at risk for. This pattern is aligned with equifinality and multifinality, in that alterations within shared developmental pathways may contribute to diverse psychopathology profiles, while related symptoms may come to rise through different neurodevelopmental pathway perturbations.

Certain limitations are noted. Firstly, youth mental health was measured through parent report. Though agreement between parent- and child-report varies^72–74^, parent-report measures examined a wider range of psychopathology across more time points and, at earlier ages, may be recommended over child-reported scoring^75^. Secondly, SOMs are sensitive to training parameters and non-deterministic generation. While SOMs are by nature exploratory, we validated our findings with robust testing of the topology, comparison to external variables and stability over time. External replication would necessarily yield a different map topology due to random initialisation and ordering of participants during training; however, future work in other samples is recommended to establish generalisability of psychopathology profiles. Finally, microarray gene expression data stemmed from 6 adult donors rather than the ABCD sample. However, an age-matched atlas of sufficient resolution is not currently available.

### Conclusions

We reconceptualised psychopathology in young adolescents as a series of dimensional topologies, extending the understanding of mental health past case-control division and retaining interpretable links between symptoms and biology at the individual level. Symptoms, genetics and connectivity converged on a hierarchy of psychopathology, including an overarching p-factor and further symptom-type axes. Both the encoding and expression of risk genes for psychopathology converged on a shared group of mechanisms including neurodevelopmental and regulatory cellular processes. Psychopathology in young adolescents emerges as a combination of non-linear symptom dimensions with biological basis in atypical development/functioning of general neurodevelopmental pathways.

## Methods

### Data

#### Participants

Participants for the study originated from the Adolescent Brain Cognitive Development (ABCD) study^76^. The ABCD is an ongoing longitudinal cohort study which initially recruited 11875 young people at ages 9-10 from across 21 study sites in the United States with the aim to follow them up during development. The study incorporates a wide battery of assessments from within neuroimaging, genetic, clinical, cognitive, mental health and environmental domains, with the goal of understanding young people’s development. Detailed sampling strategy and demographic details has been described in detail elsewhere ^77^, and the demographic data for the sample after our filtering procedures is summarised in Supplementary Materials (Table S5). For the present study, we used behavioural data for baseline (N=11862), 1-year (N=11201), 2-year (N=10897), 3-year (N=10098) and partial 4-year (N=4677) follow-up, as well as baseline resting-state fMRI (N= 4264) and polygenic risk score data (N=5672), available in the ABCD 5.1. Release.

#### Behavioural Measures

Continuous measures of mental health symptoms were used to characterise psychopathology. The Parent Child Behavior Checklist (CBCL)^42^ was used to assess symptoms of internalising, externalising, social and cognitive problems. CBCL contains 117 items grouped into 8 subscales: Rule-breaking Behaviour, Aggressive Behaviour, Withdrawn, Somatic Complaints, Anxious/Depressed, Social Problems, Thought Problems, and Attention Problems. Additionally, symptoms of mania were measured by the 10-item Parent General Behavior Inventory – Mania Scale^43^. One subscale of the Sleep Disturbance Scale for Children^44^, *Disorders of Initiating and Maintaining Sleep (DIMS)*, was also included due to the importance of sleep disturbances as a transdiagnostic symptom present across psychopathology presentations^78,79^. To complete an external validation step, we also used the parent-reported Kiddie Schedule for Affective Disorders and Schizophrenia^47^. Data from the baseline measurement were used for the main analysis, with data from the 1-year, 2-year, 3-year and partial 4-year follow-up timepoints for a stability analysis.

All described measures relied on parent-reported scoring. Participants who were missing data on complete assessments were excluded, while partially missing responses were imputed using the k-Nearest Neighbour algorithm^80^. Raw scores for each subscale were used, following recommendations to use raw over t-scored data in this sample^81^ and the lack of t-score calculations for PGBI-Mania and Sleep Disturbance Scale for Children scales. Participants were excluded from analysis if they did not have the CBCL measure, resulting in a baseline sample of 11862. As the measures of Mania and Sleep problems were scored on a different Likert range, the scores were harmonized with CBCL by transforming into equidistant scores using min-max scaling, followed by z-score transformation, projection to 0-2 range matching the CBCL subscales, and recalculation of summary scores^82^.

#### Functional Neuroimaging

Resting-state fMRI data from the baseline scan was used. 4326 scans were available after QC and processing, and participants who did not have matched psychopathology data were excluded, resulting in 4264 retained for PLS. The detailed imaging protocol for fMRI imaging in the ABCD cohort is described in Casey et al. ^76^. Minimally processed resting-state fMRI data were processed by researchers (SA, DB, SWY) at McGill University and Yale University. The preprocessing pipeline, described originally Dhamala et al. ^83^, as well as further processing steps, are included in Supplementary Materials.

After pre-processing, full Pearson correlations were computed between the time series of each pair of nodes and transformed using the Fisher-z transformation. The Shaefer-100 atlas was used for parcellating the data. For PLS analysis, per row, the top 20% of connections were retained; as this resulted in an asymmetrical matrix, an edge_ij_ was retained if the connection survived the threshold for node_i_ *or* node_j_^84^.

#### Polygenic risk scores

Genetic risk for mental health problems is polygenic, emerging from a combination of genetic variants each conferring a very small influence^85,86^. To estimate individual-level genetic liability to psychopathology, we used polygenic risk scores (PGRs). Genome-wide association studies (GWAS) conducted in a base sample elucidate loci relevant to the continuous or binary phenotype of interest, and the markers are ranked by the strength of evidence of the association.

In an independent target cohort, individuals are then genotyped and the polygenic risk score is calculated by summing up the risk alleles and weighting them by the effect sizes associated with the loci within the GWAS^87^. It is important to note PGRs can only provide a certain level of predictive accuracy, limited by the extent of genetic contribution to prediction of risk for psychopathology^88^.

For this analysis, polygenic scores were calculated (YE, VW) for 17 common mental health disorders (Attention-Deficit Hyperactivity Disorder (ADHD), Autism, Anorexia, Anxiety, Bipolar Disorder, Major Depressive Disorder, Obsessive-Compulsive Disorder (OCD), Panic Disorder, Post-traumatic Stress Disorder, Schizophrenia and Substance Use Disorder (SUD) using summary statistics with the exclusion of the UK Biobank. Scores for 5678 individuals were available, and after exclusion of participants without matched symptom scores, 5672 were retained. Polygenic score was derived using continuous shrinkage (PRS-cs), a Bayesian algorithm which infers posterior effect sizes without requiring a predetermined p-value threshold ^89^. Default parameters (Gamma priors a=1, b=0.5, shrinkage factor *φ* = 0.01) were applied. Variant-level effect-sizes were estimated for all common small nucleotide polymorphisms (SNPs) (frequency > 0.1%) present in both ABCD and HapMap3 European-ancestry reference panels, with aggregate PGRs calculated using PLINK^90^. A detailed pipeline for obtaining the PGR scores reported in this sample is described in He et al.^91^.

### Data Analysis

#### Self-organising maps

Self-organising maps (SOM) are an unsupervised machine learning data reduction method capable of representing multivariate data on an interpretable, two-dimensional plane or “map”^45,46^. In our pipeline of SOM training, an artificial neural network was initialised with a grid size of 24 x 24 nodes, calculated using the equation 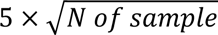 ^92^. A 10- dimensional weight vector was assigned to each node using random weights from within the sample. Then, the iterative process of SOM training proceeded as follows: i) each input vector (participant) was mapped to a node in the network to which it was closest in Euclidean distance, referred to as Best Matching Unit (BMU), ii) the weights of that BMU as well as surrounding nodes were updated with each input vector at a learning rate (0.5) and neighbourhood sigma (1.5) dependent on model parameters iii) the process is repeated for each participant (N=11862) and iterated 5000 times. Different combinations hyperparameters including learning rate, neighbourhood sigma and iteration were tested, and the winning model was established based on optimisation of quantisation and topographic errors. SOMs were trained using the MiniSom library ^93^ in Python. In the trained SOM, adjacent nodes have similar weights, resulting in continuous dimensional transitions between areas of topology. Additionally, each weight vector reflects the mental health scores of participants for whom that was their BMU, meaning that youth with similar mental health phenotypes are mapped proximally, while those dissimilar are placed further apart.

SOMs have been successfully applied in prior research to map cognition^94–97^ and psychopathology diagnoses and symptoms (e.g. stress, suicidal behaviour)^98–101^. Advantages of the method, such as the ability to map multivariate, non-linear relationships and effective handling of missing data^102–104^, make the method particularly well-suited for applications within exploring complex mental health data.

#### Psychopathology profiles

Prior approaches to parsing SOM topology include clustering the weights of the map’s nodes with algorithms such as k-means^96,100^. However, we sought out a different approach for two reasons. Firstly, as discussed prior, assignment of mental health symptoms to discrete groups often imposes an artificial separation, with studies often finding low modularity and suboptimal clustering stability^40,41^. Secondly, the SOM represents a continuous latent space where adjacent nodes take on continuous vector weights throughout training, and a strict partitioning of SOM topology based on a clustering algorithm imposes artificial boundaries on the smooth transitions between neighbouring nodes. We therefore followed a novel approach to investigating the topology of the SOM:

1. We generated groups of 500 simulated individuals matching each of 8 theory-driven psychopathology profiles, generated by extracting the covariance matrix of the original dataset and manipulating individual feature levels to simulate heightened symptoms (defined as >= 95^th^ percentile) in profile-specific combinations. This ensured symptom patterns followed feasible symptom elevations present in the normative sample. A complete list of profiles, constituent symptoms and example items from relevant measurement subscales is provided in Supplementary Table S6.
2. Data from sets of simulated individuals exhibiting a candidate profile were used to train an ensemble tree-based classifier, random forest^105^, implemented using scikit-learn in Python. The trained model was then used to classify the weight vectors of SOM neurons into one of 8 profiles or a null (median) profile. This allowed us to observe spatially contiguous regions (“zones”) on the SOM representing psychopathology profiles.
3. After assignment of nodes, permutation testing was used to establish the robustness of identified zones within SOM topology. The neuron weight vectors of the SOM were randomly shuffled 1000 times and the classifier was applied to the shuffled maps, recording the sizes of adjacent (including diagonal) nodes on the map. This allowed for the construction of a null distribution of cluster sizes expected by chance. Observed zones from the original classification were compared to the null distributions and those surpassing a threshold of five contiguous nodes were retained as non-random, statistically robust profile zones. Two profiles (*Mood, Mania*) were eliminated at this stage – *Mania* symptoms were very sparsely endorsed in the sample and the combination of symptoms within *Mood* overlapped with the *Internalising* profile.

#### Neurobiological Modelling PLS-C

Partial Least Squares (PLS) correlation^106,107^ was used to understand the association between SOM-derived psychopathology profiles and i) resting-state connectivity; and ii) polygenic risk scores for common mental health diagnoses. PLS is a data-driven technique which analyses the relationship between two datasets storing information about the same sample and maximizes covariance between the datasets. It is a technique well-suited for latent structure discovery in large-sample multivariate data^108^ and has been previously applied to investigate the association between transdiagnostic psychopathology and functional connectivity in another sample of youth^109^.

PLS analysis was implemented as follows: In our neuroimaging model, thresholded rsfMRI connectivity matrices constituted matrix **X** (participants x edgewise functional connectivity strength) and psychopathology profile based on BMU from SOM constituted **Y** (participants by one-hot encoding of membership in profile). Data was z-scored and the cross-covariance matrix was computed as R = Y^T^ × X, followed by singular value decomposition: R

= U × **Δ** × V^T^, where:

U – singular vectors of behavioural weights V – singular vectors of brain weights

**Δ** – diagonal matrix of singular values^48^.

Pairs of orthogonal latent variables (LV) can then be computed by projecting the original X and Y onto V and U, respectively:

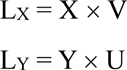

From pairs of columns in L_X_ and L_Y_, PLS exposes latent variables which represent the maximal explained covariance between X and Y, which we can quantify for each LV through the squared singular value divided by squared sum of all singular values^51^. The final expression of each latent variable, known as loading, is the correlation between participant-level scores from L_X_ and L_Y_ and the original data.

To assess the significance of derived LVs, we performed permutation testing by randomly reshuffling rows of the X matrix and recalculating the PLS model over 1000 iterations to establish a null distribution. To identify stable loadings, data was resampled with replacement and a bootstrap ratio (BSR) was calculated by dividing each loading by its standard deviation from bootstrap iterations. Resulting BSRs are interpreted similarly to Z-scores^48^ where a threshold of 1.96 corresponds to a p < 0.05 and is used as a significance mask on edge values from PLS. PLS correlation analysis was implemented using the pyls package (https://github.com/rmarkello/pyls) for Python.

Confounding variables including sex assigned at birth, age, scanner site and mean scanner motion were assessed post-hoc using a GLM to ensure identified relationships between the datasets were not attributable to these external variables.

In the polygenic risk scores model, an analogous pipeline was followed with polygenic risk scores of mental health conditions as **X** (participants x scores on 17 PGRs) and psychopathology profile based on BMU from SOM constituted **Y** (participants x one-hot encoding of membership in profile).

#### Gene Expression

Regional gene expression data were obtained from the Allen Human Brain Atlas (AHBA; https://human.brain-map.org/). The AHBA is an atlas constructed from microarray analysis validated using RNA-Seq. The sample consists of postmortem brain tissue samples from six healthy donors (5 male, aged 18-68) with no known neurological or psychiatric disorder history. Only two brain samples included the right hemisphere, so analysis was only conducted on the left hemisphere (and matched accordingly when gene expression data was combined with other analyses). Details of sample acquisition are detailed in relevant Allen

Human Brain map documentation (https://community.brain-map.org/t/documentation-human-brain-atlas), and details of data processing are available elsewhere^110^.

#### Gene Ontology and Expression

We sought to identify if the genes implicated in the polygenic risk scores PLS model and psychopathology-relevant regional gene expression patterns in the brain converge on shared biological functions. We therefore extracted two gene sets, one derived from a regional transcriptome x psychopathology-related functional connectivity contrast and one from previously identified genes conferring risk for psychopathology.

For the first gene set, we extracted patterns of gene expression covariant with psychopathology-relevant brain regions using a PLS correlation analysis. The first matrix was the expression values of 13561 genes mapped to 50 left hemisphere nodes from the AHBA. The second input matrix was the mean difference between connectomes of individuals in the “No Problems” profile and those in “High Psychopathology”, “Externalising” and “Somaticising” profiles, resulting in a vector of psychopathology-related connectivity contrasts across nodes. The PLS identified shared covariance between regional transcription and regional psychopathology-relevant connectivity contrast.

The second gene set was extracted based on genes constituting genetic risk for psychiatric diagnoses from GWAS analysis. For each PRS indicated as a stable contribution to the first LV in the PLS analysis of PRS scores (ADHD, Anxiety, Autism, Bipolar, Bipolar Type 2, MDD, PTSD, SUD), we extracted the contributing genes indicated in the original GWAS based on which the PRS was calculated (Supplementary Table S7).

Comparative pathway enrichment analysis^111^, also called over-representation or functional enrichment analysis, detects whether genes in a provided set are over- represented/enriched in particular biological pathways relative to a background set. We used multi-query analysis with two gene lists as this assesses enrichment across two sets simultaneously, producing an FDR-adjusted *p* value for each list individually as well as the size of the intersection between both, resulting in additional insight into shared enriched ontology terms and the relative contribution of each list to the overlapping biological functions. In our analysis, the two gene sets were annotated using using Gene Ontology (GO)^112^ including biological process (BP), molecular function (MF) and cellular component (CC) terms. Comparative pathway enrichment was completed using g:Profiler^113^. Results were filtered to include GO terms significantly (FDR-corrected p_adj_ < 0.05) enriched in both analysed lists.

Biological process terms were later visualised using Cytoscape v 3.10.4^114^ with EnrichmentMap v 3.5.0^115^. Functional clusters were grouped and annotated using Cytoscape’s AutoAnnotate plugin using MCL Cluster algorithm in clusterMaker2^116^. Edge similarity thresholds for clustering were adjusted for BP, CC and MF separately to optimise cluster cohesion and minimise singleton nodes (threshold BP = 0.55, threshold CC= 0.5, threshold MF = 0.5). In the presented BP annotation, four singleton clusters were removed by the plotting algorithm but included five significantly enriched shared GO terms: commissural neuron axon guidance (GO:0071679), neural precursor cell proliferation (GO:0061351), homeostatic process (GO:0042592), growth (GO:0040007) and regulation of molecular function (GO:0065009).

## Data availability

Data used in the preparation of this article were obtained from the Adolescent Brain Cognitive Development^SM^ (ABCD) Study (https://abcdstudy.org), held in the NIMH Data Archive (NDA). This is a multisite, longitudinal study designed to recruit more than 10,000 children age 9-10 and follow them over 10 years into early adulthood. The ABCD Study® is supported by the National Institutes of Health and additional federal partners under award numbers U01DA041048, U01DA050989, U01DA051016, U01DA041022, U01DA051018, U01DA051037, U01DA050987, U01DA041174, U01DA041106, U01DA041117, U01DA041028, U01DA041134, U01DA050988, U01DA051039, U01DA041156, U01DA041025, U01DA041120, U01DA051038, U01DA041148, U01DA041093, U01DA041089, U24DA041123, U24DA041147. A full list of supporters is available at https://abcdstudy.org/federal-partners.html. A listing of participating sites and a complete listing of the study investigators can be found at https://abcdstudy.org/consortium_members/. ABCD consortium investigators designed and implemented the study and/or provided data but did not necessarily participate in the analysis or writing of this report. This manuscript reflects the views of the authors and may not reflect the opinions or views of the NIH or ABCD consortium investigators.

All data files are available from the ABCD Study Data Repository from the NIMH Data Archive (https://nda.nih.gov/abcd). The DOI for this NDA Study is 10.15154/z563-zd24.

## Code availability

Code used for the analysis is available in a Github repository (https://github.com/mbj27/Topology-of-adolescent-mental-health).

## Authorship contribution

MBJ led study design, data analysis, data organization, manuscript drafting and editing. AM, KF, ET contributed to data analysis. YH, RK and SA contributed to data organisation and processing. VW, DB and SWY contributed to data organisation, data processing and manuscript editing. DEA led study design, supervision and manuscript drafting and editing.

