## Supplementary Materials for "The topology of adolescent mental health"

### Supplementary Results

#### *Connection type*

The plots in Figure 4 show the spatial distributions of the connections that map to the psychopathology topologies, but they do not tell us about the role of those connections within the brain. For that reason, investigated whether the stable (BSR-thresholded) edges implicated by the latent variables differ by *type* of connection between LV1 and LV2. We first assessed rich club organisation based on a group-averaged, binarised consensus network and calculated the nodal degree, identifying 36 rich club nodes. Connections were then classified as rich (rich club node to rich club node), feeder (rich to non-rich) and local (non-rich to non-rich)[1]. Chi-square test was used to establish significant differences in connection type across LV1, LV2 and the consensus network, revealing a significant difference in the distribution of edge types, $\chi^2(4, 2301) = 14.28$, $p = 0.006$. The local connections in LV2 emerged as most enriched ($z = 2.7$), suggesting LV2 possibly reflects variation in local connectivity patterns; however, the magnitude of the model effect was very small (Cramér's $V = 0.056$) suggesting minor differences in connection type. Further details are available in Supplementary Materials (Figure S5, Table S1).

#### *Stability across measurement time points*

We next tested whether participants' assignment to psychopathology profiles was stable across measurement time points. To investigate this, we calculated the centroid of each psychopathology profile by averaging the weight vectors of SOM nodes classified to the profile. We then recalculated each individual's BMU at every measurement time point (baseline, 1-year, 2-year, 3-year and 4-year follow-up) and derived the Euclidean distance in feature space between their BMU weight vector and each profiles' centroid vector. We finally correlated each participant's distance to each centroid at baseline to their distances at followups. This would mean that a participant who remained in the same place on the map over time would have a correlation of 1, and if their placement at follow-up had no correspondence to baseline, they would have a correlation of 0. The majority of participants exhibited stability across measurement time points inferred from the high frequency of high correlation coefficients.

### Supplementary Results: Figures

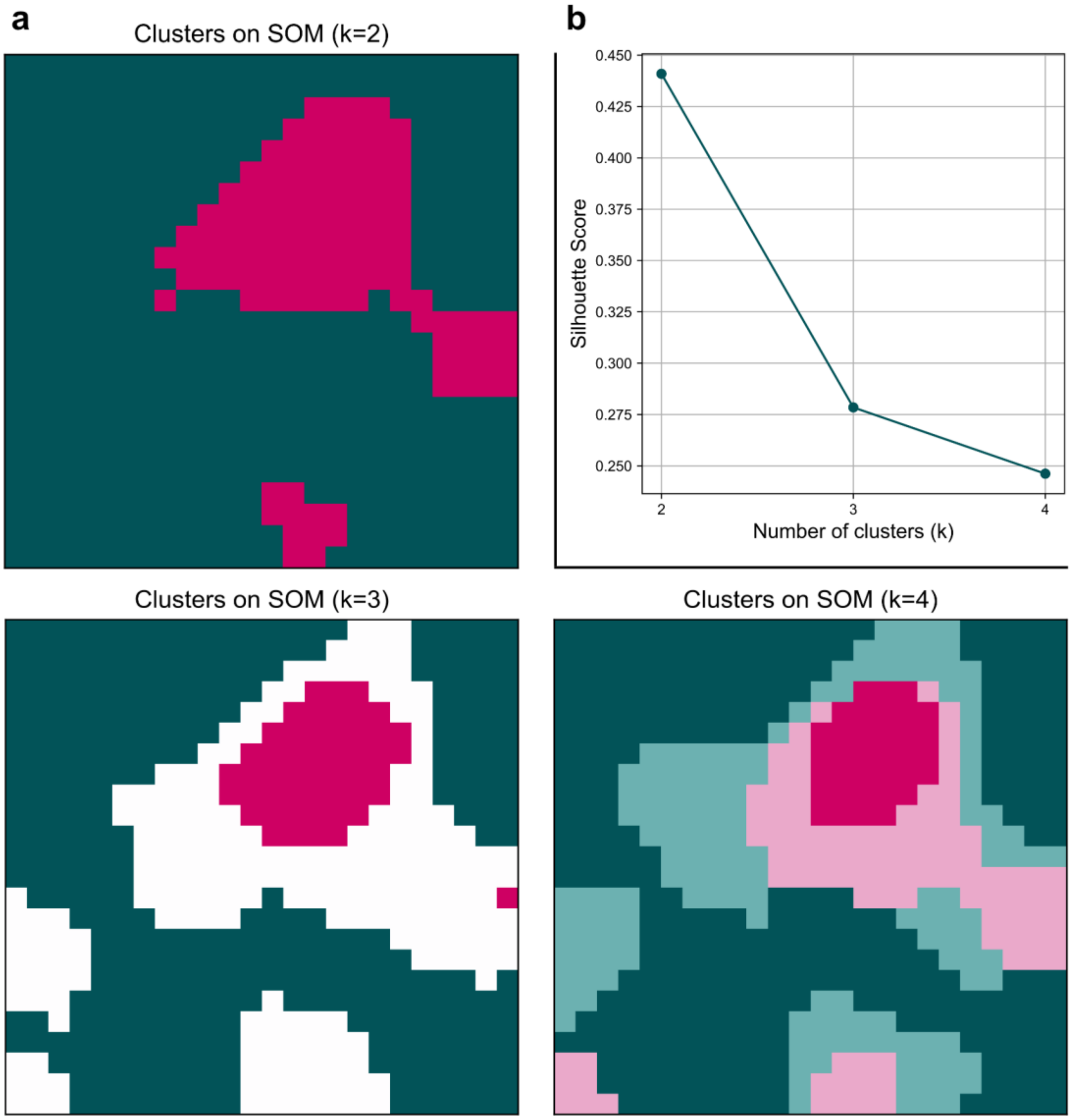


**Figure S1.**

*Limitations of discrete clustering – arbitrary clustering boundaries on continuous map.*

Panel (a) shows k-means clustering solutions of SOM weights for k=2-4. The k=2 solution broadly divides low and high weights, corresponding to lowest and highest symptom loadings. Further subdivisions artificially subdivide the continuous space into non-contiguous regions without offering interpretation of map topology. Panel (b) displays averaged silhouette scores for k=2-4 solutions, demonstrating low silhouette scores indicative of poor cluster separation.

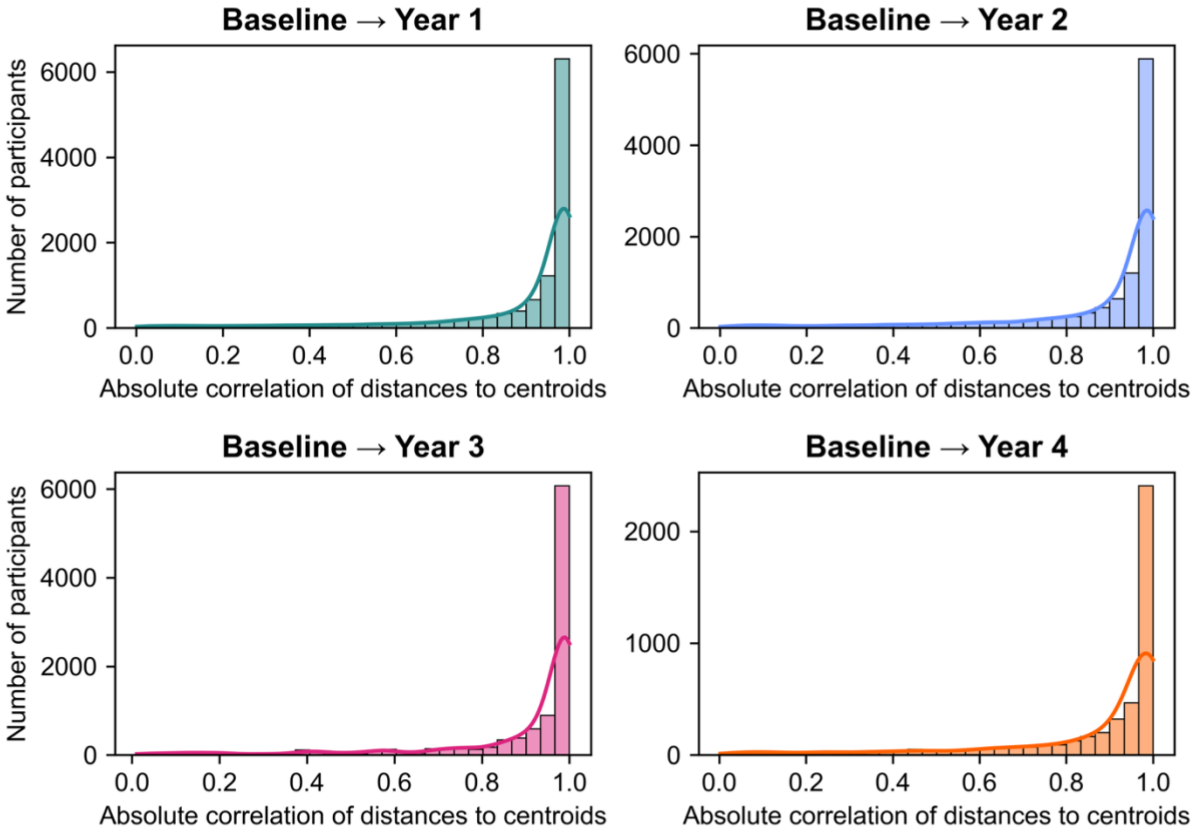


**Figure S2.**

*Distribution of participant-wise correlation of distance to psychopathology profile centroids in feature weight space between baseline measurement and follow-up time points.*

As a measure of relative stability of psychopathology profiles over time, Euclidean distance in feature weight space between a participant's BMU vector weight and the centroid of each profile was correlated between baseline and follow up time points of 1-year, 2-year, 3-year and partial 4-year follow up. Majority of participants showed high correlation in their proximity to profile centroids across time, demonstrating relative stability of psychopathology phenotype over time.

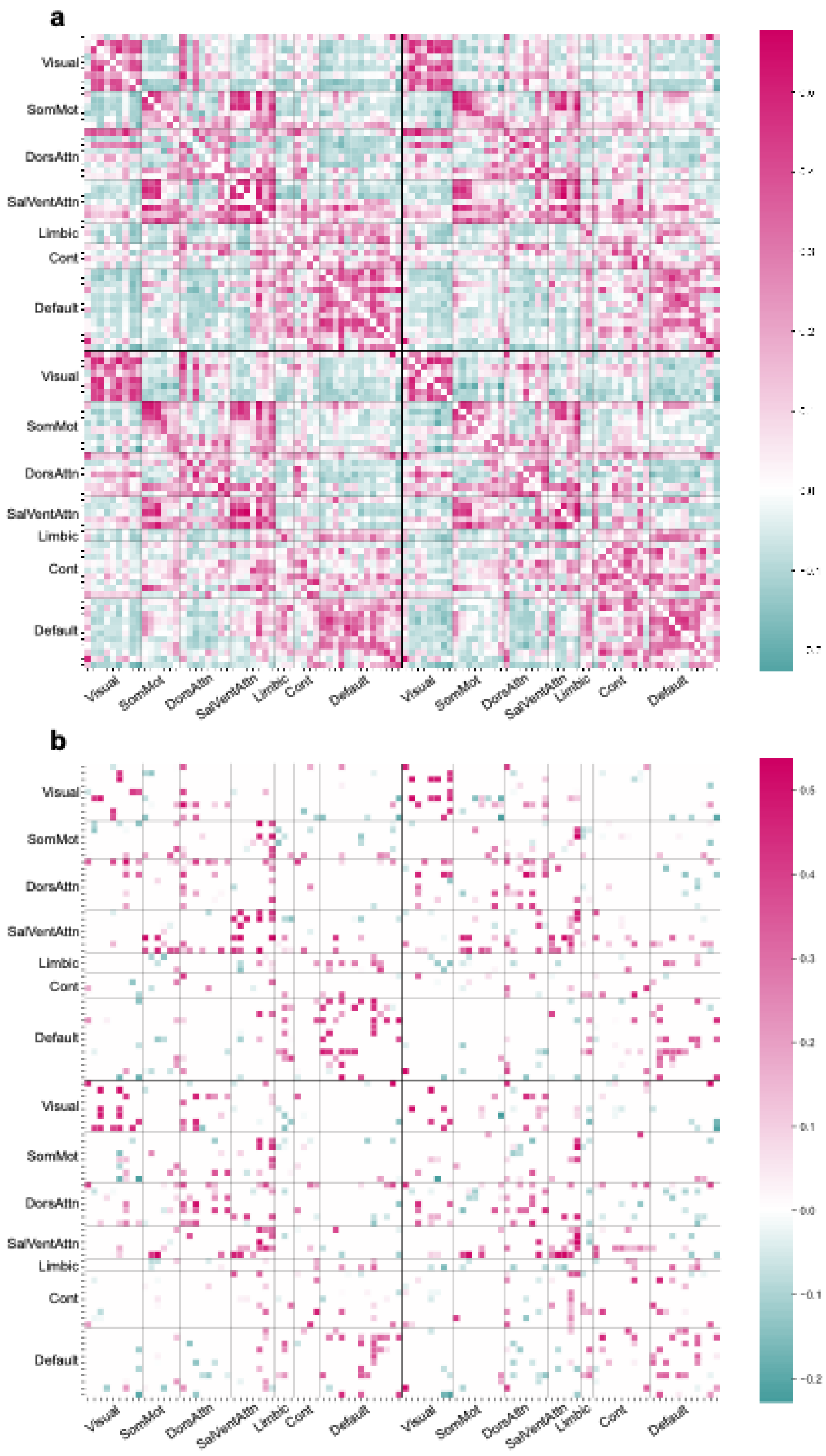


**Figure S2.**

*PLS edge loading matrixes for first significant latent variable (LV1).*

Panel A shows the unthresholded loading matrix while panel B displays the matrix thresholded by BSR > 1.96.

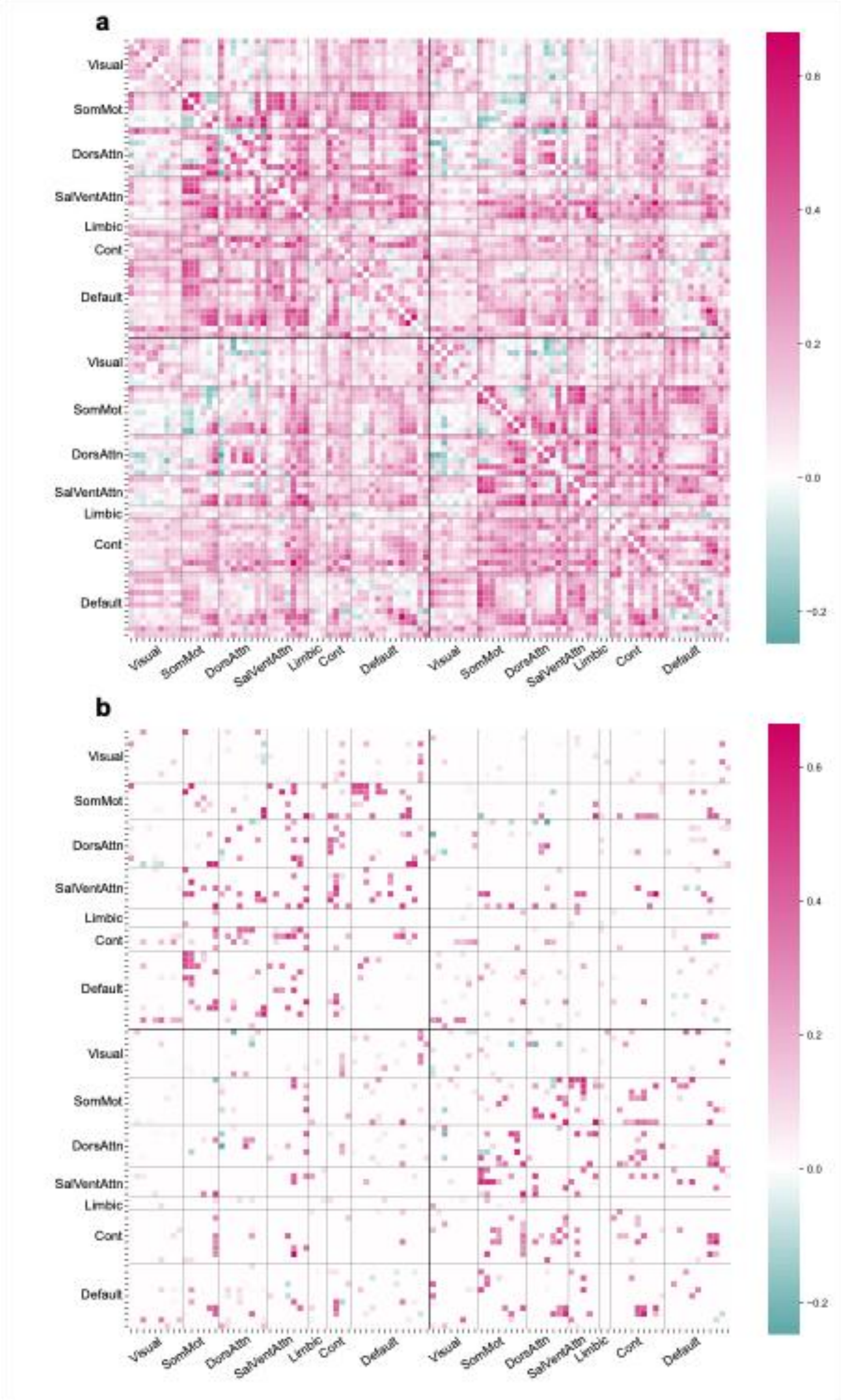


**Figure S3.**

*PLS edge loading matrixes for second significant latent variable (LV2).*

Panel A shows the unthresholded loading matrix while panel B displays the matrix thresholded by BSR > 1.96.

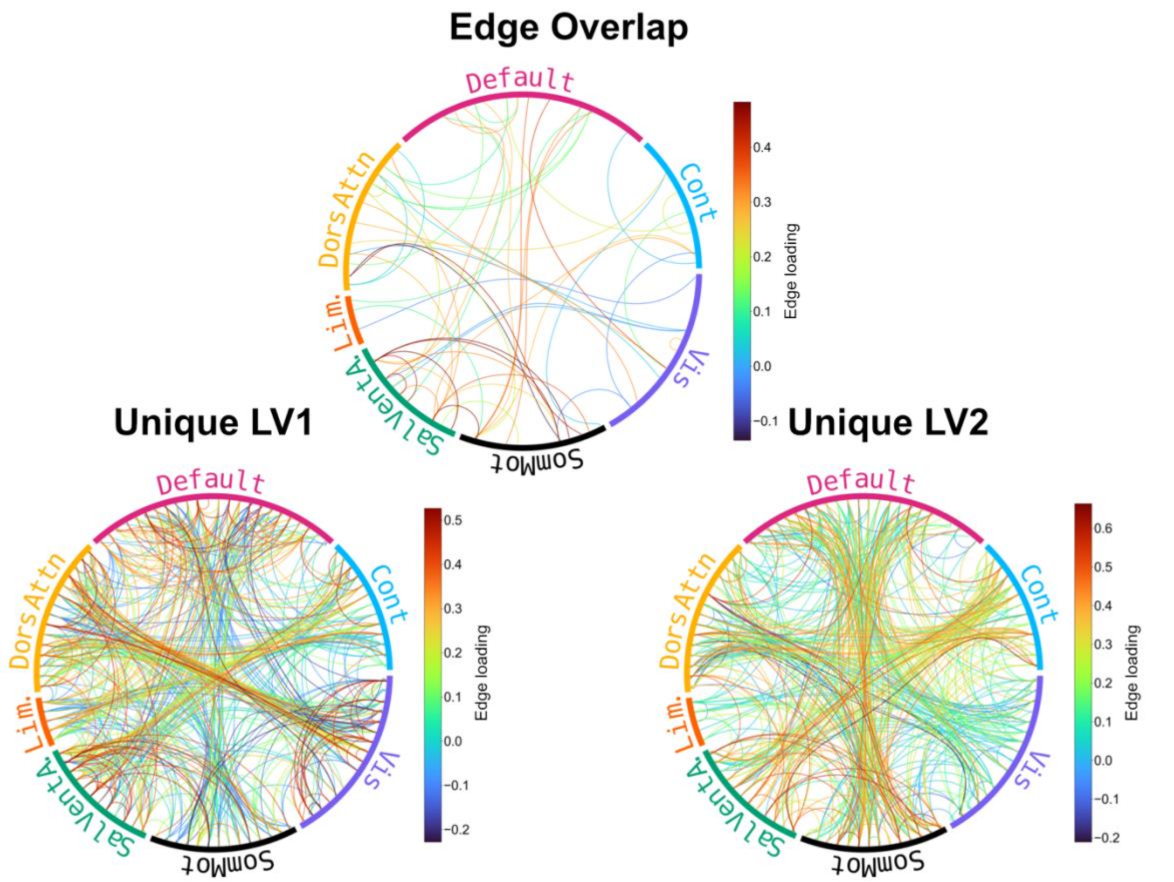


**Figure S4.**

*Shared and unique edges of LV1 and LV2 loadings on brain connectivity edges, divided by canonical network.*

Despite significant overlap in indicated canonical networks, only a small subsample of edges from rsFC PLS was shared between LV1 and LV2. In LV1, positive edge loadings represent edges associated with low symptoms, while negative edges express the pattern of higher psychopathology symptoms (p-factor). In LV2, positive edges correspond to higher social and cognitive difficulties, and negative are associated with higher internalising and somaticising problems.

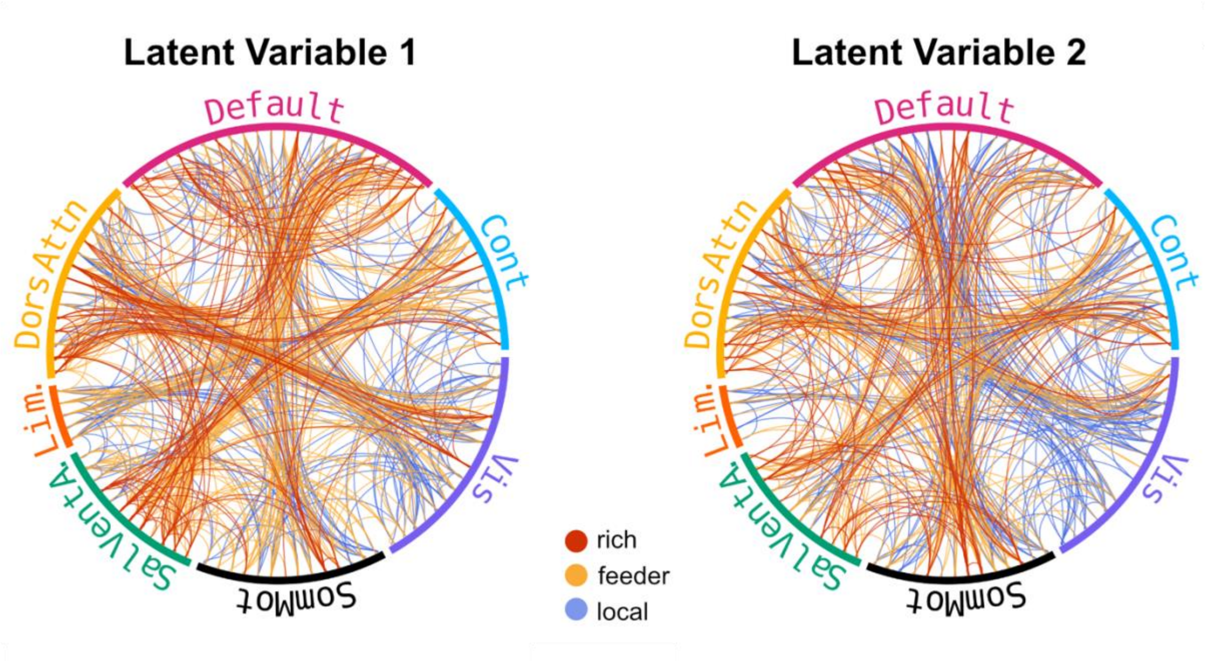


**Figure S5.**

*Rich club, feeder and local connections across LV1 and LV2*

Connections from LV1 and LV2 were then classified as rich (rich club node to rich club node), feeder (rich to non-rich) and local (non-rich to non-rich). A Chi-squared test found a significant difference in the distribution of edge types, $\chi^2(4, 2301) = 14.28$, $p = 0.006$. The local connections in LV2 emerged as most enriched ($z = 2.7$), the magnitude of the model effect was very small (Cramér's V = 0.056).

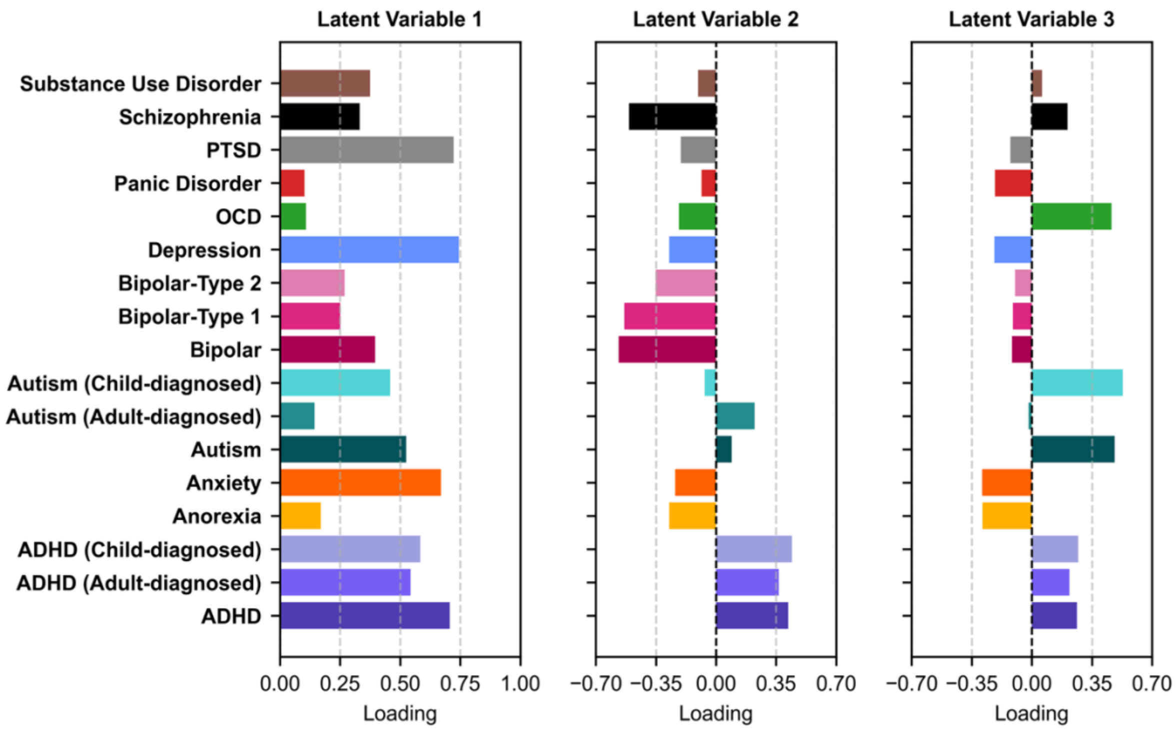


**Figure S6.**

*Significant Latent Variables from residualised PGR-psychopathology PLS*
To account for potential effects of ancestry or population stratification in polygenic risk scores, polygenic risk scores were regressed against the top 20 genetic principal components (PCs) using linear regression. A repetition of PGR-psychopathology PLS analysis using residualised PGRs revealed analogous pattern to raw values, suggesting the loadings reveal true covariance rather than shared genetic structure.

### Supplementary Results: Tables

| Group | Rich (observed/ expected) | Feeder (observed/ expected) | Local (observed/ expected) | $\chi^2$ | *p* | Cramér's V |
|---|---|---|---|---|---|---|
| LV1 | 115 / 112.7 | 256 / 253.2 | 174 / 179.1 | | | |
| LV2 | 89 / 95.0 | 186 / 213.2 | 184 / 150.8 | | | |
| Consensus | 272 / 268.3 | 627 / 602.6 | 398 / 426.1 | 14.279 | 0.006 | 0.056 |

**Table S1.**

*Distribution of connection types across LV1, LV2 and the consensus network.*

The table presents the observed and expected counts of connection types (rich, feeder, local) and the results of Pearson's Chi-square test. Cells with observed > expected are marked in green, and cells with expected > observed are marked in pink.

| Predictor | beta | HC standard error | Z value | p value |
|---|---|---|---|---|
| *intercept* | -0.03352 | 0.152488 | -0.21979 | 0.826033 |
| *interview_age* | -0.02408 | 0.019686 | -1.22325 | 0.221236 |
| *rsfmri_meanmotion* | -0.02064 | 0.019691 | -1.04842 | 0.294447 |
| ***x_score_lv1*** | **0.032774** | **0.002889** | **11.34365** | **7.97E-30*****|
| ***sex_2.0*** | **0.116363** | **0.038717** | **3.005493** | **0.002652****|
| ***sex_3.0*** | **0.685602** | **0.086359** | **7.939019** | **2.04E-15*****|
| *site_id_l_site02* | -0.01333 | 0.170661 | -0.07812 | 0.937731 |
| *site_id_l_site03* | -0.33533 | 0.175954 | -1.90581 | 0.056675 |
| *site_id_l_site04* | -0.23311 | 0.17127 | -1.36105 | 0.173498 |
| *site_id_l_site05* | 0.042731 | 0.17895 | 0.238786 | 0.811272 |
| *site_id_l_site06* | -0.10417 | 0.17314 | -0.60164 | 0.547414 |
| *site_id_l_site07* | -0.16679 | 0.195069 | -0.85502 | 0.392541 |
| *site_id_l_site08* | -0.17094 | 0.199564 | -0.85659 | 0.391674 |
| *site_id_l_site09* | 0.179683 | 0.16957 | 1.059639 | 0.289309 |
| *site_id_l_site10* | 0.246162 | 0.164452 | 1.496865 | 0.134428 |
| *site_id_l_site11* | 0.148168 | 0.175507 | 0.844225 | 0.398544 |
| *site_id_l_site12* | -0.13174 | 0.177178 | -0.74352 | 0.457165 |
| *site_id_l_site13* | 0.010357 | 0.169048 | 0.061264 | 0.951149 |
| *site_id_l_site14* | 0.221902 | 0.164243 | 1.351055 | 0.176678 |
| *site_id_l_site15* | -0.16999 | 0.194977 | -0.87186 | 0.383284 |
| *site_id_l_site16* | -0.19616 | 0.163043 | -1.20314 | 0.228923 |
| *site_id_l_site17* | 0.021735 | 0.172073 | 0.12631 | 0.899486 |
| *site_id_l_site18* | 0.184411 | 0.179246 | 1.028812 | 0.303568 |
| *site_id_l_site19* | 0.231645 | 0.174127 | 1.33032 | 0.183413 |
| *site_id_l_site20* | 0.008567 | 0.173004 | 0.049519 | 0.960506 |
| *site_id_l_site21* | -0.02595 | 0.175835 | -0.14761 | 0.882652 |
| *site_id_l_site22* | 0.082574 | 0.33788 | 0.244389 | 0.806929 |

**Table S2.**
*Post-hoc GLM covariate analysis for LV1 in FC-psychopathology PLS.*
*Note. Significant predictors are marked in bold.*

| Predictor | beta | HC standard error | Z value | p value |
|---|---|---|---|---|
| *intercept* | -0.02048 | 0.085586 | -0.23932 | 0.810858 |
| *interview_age* | -0.0003 | 0.015382 | -0.01936 | 0.984556 |
| ***rsfmri_meanmotion*** | **-0.04811** | **0.018695** | **-2.57353** | **0.010067*** |
| ***x_score_lv2*** | **0.023228** | **0.003022** | **7.686807** | **1.51E-14*****|
| ***sex_2.0*** | **-0.0858** | **0.030852** | **-2.78107** | **0.005418****|
| *sex_3.0* | 0.006199 | 0.062708 | 0.098848 | 0.921259 |
| *site_id_l_site02* | -0.01082 | 0.100414 | -0.10772 | 0.914219 |
| *site_id_l_site03* | 0.10995 | 0.112264 | 0.979384 | 0.32739 |
| *site_id_l_site04* | 0.156955 | 0.107709 | 1.457208 | 0.145059 |
| *site_id_l_site05* | -0.031 | 0.100737 | -0.30774 | 0.758279 |
| *site_id_l_site06* | -0.066 | 0.10524 | -0.62717 | 0.530546 |
| *site_id_l_site07* | 0.109825 | 0.117742 | 0.932762 | 0.350943 |
| *site_id_l_site08* | 0.181206 | 0.145299 | 1.247124 | 0.212352 |

| | | | | |
|---|---|---|---|---|
| *site_id_l_site09* | 0.048823 | 0.10014 | 0.487552 | 0.625867 |
| *site_id_l_site10* | 0.044667 | 0.094278 | 0.473783 | 0.635655 |
| *site_id_l_site11* | -0.03759 | 0.103762 | -0.3623 | 0.717131 |
| *site_id_l_site12* | 0.197364 | 0.120662 | 1.635671 | 0.101909 |
| *site_id_l_site13* | -0.02978 | 0.09439 | -0.31551 | 0.752375 |
| *site_id_l_site14* | -0.0392 | 0.097566 | -0.40177 | 0.687856 |
| *site_id_l_site15* | 0.15105 | 0.126436 | 1.194673 | 0.232215 |
| *site_id_l_site16* | 0.09004 | 0.100186 | 0.898728 | 0.368797 |
| *site_id_l_site17* | 0.095805 | 0.112071 | 0.85486 | 0.392629 |
| *site_id_l_site18* | 0.040776 | 0.114597 | 0.35582 | 0.721976 |
| *site_id_l_site19* | 0.19773 | 0.12277 | 1.610581 | 0.107271 |
| *site_id_l_site20* | 0.051927 | 0.099179 | 0.52357 | 0.600578 |
| *site_id_l_site21* | 0.08983 | 0.112925 | 0.795486 | 0.426331 |
| *site_id_l_site22* | -0.27509 | 0.176908 | -1.55501 | 0.119943 |

**Table S3.**
*Post-hoc GLM covariate analysis for LV2 in FC-psychopathology PLS.*
*Note. Significant predictors are marked in bold.*

| Predictor | beta | HC standard error | Z value | p value |
|---|---|---|---|---|
| *intercept* | 0.35280263 | 0.18687294 | 1.88792786 | 0.05903564 |
| ***interview_age*** | **0.04417142** | **0.0173163** | **2.55085702** | **0.01074584** |
| ***rsfmri_meanmotion*** | **0.10277718** | **0.02129134** | **4.82718118** | **1.38E-06** |
| ***x_score_lv1*** | **0.07432689** | **0.00920196** | **8.07728719** | **6.62E-16** |
| ***sex_2.0*** | **-0.0772134** | **0.0332225** | **-2.3241289** | **0.02011859** |
| *site_id_l_site02* | -0.3596113 | 0.19731989 | -1.8224786 | 0.0683824 |
| *site_id_l_site03* | -0.3113064 | 0.2391987 | -1.3014553 | 0.19310267 |
| *site_id_l_site04* | -0.217398 | 0.19911812 | -1.0918041 | 0.27491922 |
| ***site_id_l_site05*** | **-0.5093509** | **0.20141518** | **-2.5288605** | **0.01144335** |
| *site_id_l_site06* | -0.2512473 | 0.19696257 | -1.2756094 | 0.20209363 |
| *site_id_l_site07* | -0.2558391 | 0.21003871 | -1.2180567 | 0.22320243 |
| *site_id_l_site08* | -0.1985098 | 0.22256065 | -0.8919356 | 0.37242747 |
| ***site_id_l_site09*** | **-0.5115605** | **0.20824117** | **-2.456577** | **0.01402677** |
| ***site_id_l_site10*** | **-0.413822** | **0.2098307** | **-1.9721709** | **0.0485901** |
| *site_id_l_site11* | -0.2205324 | 0.20725457 | -1.0640652 | 0.28729917 |
| *site_id_l_site12* | -0.1206021 | 0.2070789 | -0.582397 | 0.56029932 |
| *site_id_l_site13* | -0.3488609 | 0.19642552 | -1.7760468 | 0.07572521 |
| ***site_id_l_site14*** | **-0.5677657** | **0.1922563** | **-2.953171** | **0.00314528** |
| *site_id_l_site15* | -0.0864325 | 0.22581631 | -0.3827559 | 0.70190079 |
| *site_id_l_site16* | -0.1293206 | 0.19212158 | -0.6731185 | 0.50087189 |
| *site_id_l_site17* | -0.333291 | 0.19522771 | -1.7071912 | 0.08778653 |
| ***site_id_l_site18*** | **-0.3961117** | **0.20097122** | **-1.9709871** | **0.04872535** |
| ***site_id_l_site19*** | **-0.5722374** | **0.19631716** | **-2.9148619** | **0.00355846** |

| | | | | |
|---|---|---|---|---|
| *site_id_l_site20* | -0.3418879 | 0.19884977 | -1.7193278 | 0.08555469 |
| ***site_id_l_site21*** | **-0.4245214** | **0.19952685** | **-2.1276403** | **0.03336691** |
| *site_id_l_site22* | -0.6500574 | 0.2939884 | -2.2111669 | 0.02702428 |

**Table S4.**

*Post-hoc GLM covariate analysis for LV1 in PGR-psychopathology PLS.*
*Note. Significant predictors are marked in bold.*

| Predictor | beta | HC standard error | Z value | p value |
|---|---|---|---|---|
| *intercept* | -0.1126433 | 0.15358724 | -0.7334155 | 0.46330505 |
| *interview_age* | -0.0037 | 0.01463826 | -0.252761 | 0.80045292 |
| *rsfmri_meanmotion* | 0.02962005 | 0.01590122 | 1.86275333 | 0.06249698 |
| ***x_score_lv2*** | **0.05791264** | **0.0100894** | **5.73994794** | **9.47E-09** |
| ***sex_2.0*** | **-0.119791** | **0.02890835** | **-4.1438201** | **3.42E-05** |
| *site_id_l_site02* | 0.17198551 | 0.16191611 | 1.06218901 | 0.28814989 |
| *site_id_l_site03* | 0.23334747 | 0.1883143 | 1.23913834 | 0.21529427 |
| *site_id_l_site04* | 0.20615663 | 0.16163872 | 1.2754161 | 0.202162 |
| ***site_id_l_site05*** | **0.34722665** | **0.16715845** | **2.0772306** | **0.03778028** |
| *site_id_l_site06* | 0.09394322 | 0.16535383 | 0.5681345 | 0.56994364 |
| *site_id_l_site07* | 0.25155913 | 0.17820203 | 1.41165134 | 0.15805265 |
| *site_id_l_site08* | 0.08407189 | 0.18501009 | 0.45441787 | 0.64952809 |
| ***site_id_l_site09*** | **0.41380516** | **0.17199292** | **2.40594297** | **0.01613078** |
| *site_id_l_site10* | 0.28713263 | 0.16964853 | 1.69251472 | 0.09054787 |
| *site_id_l_site11* | 0.12563531 | 0.17019632 | 0.73817876 | 0.46040583 |
| *site_id_l_site12* | 0.07907963 | 0.17791867 | 0.44447064 | 0.65670235 |
| *site_id_l_site13* | 0.13362191 | 0.1627672 | 0.82093879 | 0.41168114 |
| *site_id_l_site14* | 0.2362083 | 0.15762983 | 1.49849993 | 0.13400341 |
| *site_id_l_site15* | 0.05239143 | 0.18499929 | 0.28319798 | 0.77702508 |
| *site_id_l_site16* | 0.09204865 | 0.1584876 | 0.58079403 | 0.56137928 |
| *site_id_l_site17* | 0.10494213 | 0.16197695 | 0.64788314 | 0.51706054 |
| *site_id_l_site18* | 0.1716213 | 0.16972133 | 1.01119463 | 0.31192328 |
| *site_id_l_site19* | 0.10923019 | 0.16383728 | 0.66669925 | 0.50496426 |
| *site_id_l_site20* | 0.1728872 | 0.16251387 | 1.06383045 | 0.28740551 |
| *site_id_l_site21* | 0.24782196 | 0.16534164 | 1.49884781 | 0.13391312 |
| *site_id_l_site22* | 0.32310787 | 0.19796543 | 1.63214284 | 0.1026494 |

**Table S5.**

*Post-hoc GLM covariate analysis for LV2 in PGR-psychopathology PLS.*
*Note. Significant predictors are marked in bold.*

| Predictor | beta | HC standard error | Z value | p value |
|---|---|---|---|---|
| *intercept* | 0.350233039 | 0.190721687 | 1.836356658 | 0.066304925 |
| *interview_age* | -0.014924016 | 0.014575994 | -1.02387642 | 0.305893649 |
| *rsfmri_meanmotion* | 0.024220937 | 0.019277887 | 1.256410386 | 0.208967218 |
| ***x_score_lv3*** | **0.078633674** | **0.014269551** | **5.510592185** | **3.58E-08** |
| *sex_2.0* | -0.027612498 | 0.028019678 | -0.985468093 | 0.324394197 |
| *site_id_l_site02* | -0.357925799 | 0.197342919 | -1.813725066 | 0.069720064 |
| *site_id_l_site03* | -0.343034661 | 0.223501552 | -1.534820036 | 0.12482804 |
| *site_id_l_site04* | -0.376497149 | 0.19820069 | -1.899575369 | 0.057488867 |
| *site_id_l_site05* | -0.343887781 | 0.202574857 | -1.697583728 | 0.089586356 |
| *site_id_l_site06* | -0.280248829 | 0.198807134 | -1.409651774 | 0.158642531 |
| *site_id_l_site07* | -0.334539493 | 0.208318572 | -1.605903351 | 0.108295147 |
| *site_id_l_site08* | -0.149184364 | 0.22572491 | -0.660912275 | 0.508668574 |
| *site_id_l_site09* | -0.380077737 | 0.206218022 | -1.843086908 | 0.065316323 |
| *site_id_l_site10* | -0.364717628 | 0.208928517 | -1.745657477 | 0.080870488 |
| ***site_id_l_site11*** | **-0.422983382** | **0.203045442** | **-2.083195658** | **0.037233393** |
| *site_id_l_site12* | -0.271698408 | 0.207183662 | -1.311389158 | 0.18972632 |
| *site_id_l_site13* | -0.341168957 | 0.197490769 | -1.727518498 | 0.084074586 |
| *site_id_l_site14* | -0.387562062 | 0.194595071 | -1.991633493 | 0.046411286 |
| *site_id_l_site15* | -0.319380122 | 0.219474466 | -1.455204005 | 0.145612792 |
| *site_id_l_site16* | -0.370599898 | 0.193933172 | -1.910967027 | 0.056008818 |
| *site_id_l_site17* | -0.373343592 | 0.195946049 | -1.905338705 | 0.056736057 |
| *site_id_l_site18* | -0.340021519 | 0.20155504 | -1.686990904 | 0.091605101 |
| *site_id_l_site19* | -0.295240546 | 0.199880573 | -1.477084746 | 0.139652922 |
| *site_id_l_site20* | -0.288721467 | 0.200254158 | -1.441775142 | 0.149365818 |
| *site_id_l_site21* | -0.280801604 | 0.202150557 | -1.389071634 | 0.164810968 |
| *site_id_l_site22* | -0.249913283 | 0.300385266 | -0.831975837 | 0.405422583 |

**Table S6.**
*Post-hoc GLM covariate analysis for LV3 in PGR-psychopathology PLS.*
*Note. Significant predictors are marked in bold.*

### Supplementary Methods

#### *Neuroimaging preprocessing pipeline*

Minimally processed resting-state fMRI data, obtained from the ABCD NIMH Data Archive, were preprocessed by researchers (SA, DB, SWY) at McGill University and Yale University. Further preprocessing was carried out according to the pipeline described in Dhamala et al. [2] and included removal of 4 initial frames, alignment to T1-weighted scans using boundary-registration, filtering of respiratory pseudo-motion using a band-stop filter of 0.31-0.43 Hz and calculation of framewise displacement (FD) and DVARS. Frames exceeding FD>0.3mm or DVARS>50 were flagged along with one preceding and two succeeding frames, and excluded if fewer than 500 frames remained, if more than half frames were censored or if any frame exceeded FD>0.5mm. In non-censored frames, nuisance regression removed 18 covariates: global signal, six motion parameters, mean ventricular signal, mean white matter signal, and their temporal derivatives, estimating regression coefficients from non-censored volumes. Motion outliers were scrubbed and interpolated across censored frames using least squares spectral estimation, and the timeseries was band-pass filtered at 0.008-0.09 Functional connectivity was computed using Pearson correlation and transformed using the Fisher-z transformation. Finally, runs were averaged within each participant for the same measurement year.

**Supplementary Methods: Tables**

| | CBCL sample | | PGS sample | | Imaging sample | |
|---|---|---|---|---|---|---|
| **Characteristic** | **Mean** | **SD** | **Mean** | **SD** | **Mean** | **SD** |
| **Age (months)** | 118.98 | 7.5 | 119.24 | 7.51 | 119.23 | 7.5 |
| **Sex** | **N** | **%** | **N** | **%** | **N** | **%** |
| **Male** | 6185 | 52.141 | 3011 | 53.085 | 2188 | 51.313 |
| **Female** | 5674 | 47.833 | 2661 | 46.915 | 2075 | 48.663 |
| **Intersex** | 3 | 0.025 | 0 | 0.000 | 1 | 0.023 |
| **Race** | | | | | | |
| **White** | 7506 | 63.278 | 5457 | 96.209 | 2794 | 65.525 |
| **Black/African American** | 1866 | 15.731 | 1 | 0.018 | 584 | 13.696 |
| **Mixed Race** | 1479 | 12.468 | 174 | 3.068 | 545 | 12.781 |
| **Asian** | 243 | 2.049 | 0 | 0.000 | 81 | 1.900 |
| **Native American/ Alaska Native** | 62 | 0.523 | 16 | 0.282 | 23 | 0.539 |
| **Pacific Islander** | 15 | 0.126 | 0 | 0.000 | 7 | 0.164 |
| **Other/Not reported** | 691 | 5.825 | 24 | 0.423 | 230 | 5.394 |
| ***Sum*** | *11862* | *100* | *5672* | *100* | *4264* | *100* |

**Table S5.**
*Demographic details of baseline sample from ABCD cohort.*

| ***Psychopathology Profile*** | ***Component Subscales*** *Example items* | | |
|---|---|---|---|
| ***Internalising*** | **Anxious/ depressed** | **Withdrawn/ depressed** | **Somatic Problems** |
| | *Unhappy, sad, or depressed* | *Would rather be alone than with others* | *Aches or pains* |
| | *Too fearful or anxious* | *Withdrawn, not involved with others* | *Nausea, feels sick* |
| ***Externalising*** | **Rule breaking** | **Aggression** | |
| | *Lying or cheating* | *Gets in many fights* | |
| | *Steals outside the home* | *Cruelty, bullying, or meanness to others* | |
| ***Cognitive*** | **Attention Problems** | **Thought Problems** | |
| | *Can't concentrate, can't pay attention for long* | *Can't get their mind off certain thoughts* | |
| | *Impulsive or acts without thinking* | *Repeats certain acts over and over* | |
| ***Somaticising*** | **Somatic** | **Sleep** | |
| | *Aches or pains* | *The child has difficulty getting to sleep at night.* | |
| | *Nausea, feels sick* | *The child wakes up more than twice per night.* | |
| ***Social Problems*** | **Withdrawn/ depressed** | **Social Problems** | |
| | *Would rather be alone than with others* | *Not liked by other kids* | |
| | *Withdrawn, doesn't get involved with others* | *Clings to adults or too dependent* | |
| ***Mood*** | **Anxious/ depressed** | **Withdrawn/ depressed** | **Somatic Problems** |
| | *Unhappy, sad, or depressed* | *Would rather be alone than with others* | *Aches or pains* |
| | *Too fearful or anxious* | *Withdrawn, not involved with others* | *Nausea, feels sick* |
| | **Thought Problems** | **Sleep Problems** | **Mania** |
| | *Can't get their mind off certain thoughts* | *Difficulty getting to sleep at night.* | *Mood or energy shifted rapidly back and forth from happy to sad or high to low* |
| | *Repeats certain acts over and over* | *Child wakes up more than twice per night* | *Feelings or energy are generally up or down, but rarely in the middle* |

| | | |
|---|---|---|
| ***Mania*** | **Thought** | **Mania** |
| | *Can't get their mind off certain thoughts* | *Mood or energy shifted rapidly back and forth from happy to sad or high to low* |
| | *Repeats certain acts over and over* | *Feelings or energy are generally up or down, but rarely in the middle* |
| ***High Psychopathology*** | **All Symptom Subscales** | |
| ***No Problems*** | **Median profile (Normative – none or very low symptoms)** | |

**Table S6.**

*Psychopathology profiles with component symptoms and example items from relevant measurement subscales.*

*Note. CBCL Subscales: Anxious/Depressed, Withdrawn/Depressed, Somatic Problems, Social Problems, Thought Problems, Attention Problems, Rule Breaking, Aggression; GBI-Mania Scale: Mania; ABCD Sleep Disturbance Scale for Children (SDS) – Disorders of Initiating and Maintaining Sleep Subscale: Sleep Problems.*

| PGR from PLS analysis | GWAS year | DOI | Genes |
|---|---|---|---|
| ADHD[3] | 2022 | https://doi.org/10.1038/s41588-022-01285-8 | PPIH, C1orf210, ELOVL1, MED8, SZT2, HYI, PTPRF, ST3GAL3, ARTN, B4GALT2, KAT2B, FAM198A, SNRK, ANO10, ABHD5, ZNF852, ZNF660, ZNF197, GPX1, RHOA, TCTA, AMT, MST1, RNF123, AMIGO3, GMPPB, FAM212A, TRAIP, CAMKV, MST1R, CTD-2330K9.3, MON1A, RBM6, GNAT1, GNAI2, HYAL3, MAPKAPK3, IQCF3, FOXP1, VGLL3, ZNF827, LSM6, RP11-6L6.2, SH3RF2, COL19A1, FOXP2, FAM83H, FAM203A, MROH1, VPS28, CYHR1, KIFC2, PPP1R16A, MFSD3, RECQL4, LRRC14, LRRC24, C8orf82, ARHGAP39, ZNF251, ZNF34, RPL8, ZNF517, ZNF7, COMMD5, ZNF16, SORCS3, POC1B, VRK1, CDH8, TMEM200C, L3MBTL4, ARHGAP28, DCC, XRN2, NKX2-4 |
| Anxiety[4] | 2024 | https://doi.org/10.1038/s41588-024-01908-2 | NEGR1, LINC01360, WDPCP, LOC100287010, CNTNAP5, TEX41, SATB1-AS1, TOPAZ1, NEK4, ITIH1, ITIH3, ITIH4, ITIH4-AS1, SUCLG2-AS1, TAFA1, FBXL17, PRR16, GPM6A, BTN3A2, BTN2A2, BTN3A1, HIVEP2, TMEM106B, PCLO, CYP7B1, LINC00051, TSNARE1, ZDHHC21, PTCH1, LOC100507346, SORCS3, SORCS3-AS1, ELP4, PAX6, PAX6-AS1, PAUPAR, MIR130A, YPEL4, CLP1, ZDHHC5, MED19, TMX2, TMX2-CTNND1, TMEM258, MIR611, FEN1,FADS1, MIR1908, FADS2, LOC101928847, NCAM1, IGSF9B, SOX5, SP7, SP1, AMHR2, LOC101929058, CIT, LINC00333, STK24, STK24-AS1, LRFN5, FCF1, YLPM1, IFT43, GPATCH2L, VAMP2, TMEM107, SNORD118, MIR4521, BORCS6, AURKB, LINC00324, CTC1, PFAS, ASXL3, RAB27B, MMP9, SLC12A5-AS1, SLC12A5, NCOA5, BRWD1, BRWD1-AS2, BRWD1-AS1, HMGN1, GET1, WRB-SH3BGR, SDC4P, SEC14L4, SEC14L6, GAL3ST1, PES1, RBX1, SNORD140 |
| Autism[5] | 2019 | https://doi.org/10.1038/s41588-019-0344-8 | SOX7, RP1L1, RN7SKP32, RNU6-819P, ADCY8, ASAP1, MIR1302-7, C8orf90, RSU1, RNU6-543P, EGR2, RPL7AP50, MRPL35P2, GUCY1A2, PCDH9, ESRRB, SERPINA1, SERPINA2, LINC02210-CRHR1, MAPT, KANSL1, GALNT1, NEDD4L, ZNF568, MACROD2, KIZ, XRN2, ZNF877P, RN7SL831P, EEF1A1P11, LINC01776, SNX7-DT, THAP3P1, CDK4P1, RPL23AP22, LINC01031, PKP4, SGO1-AS1, CADPS, RSRC1, RN7SKP222, FGF12, Y_RNA, LINC02994, MIR9-2HG, LINC02058, NIHCOLE, RNU6-334P, KCNN2, GALNT10, ADTRP, SPTLC1P2, RNU6-1060P, ISG20L2P1, SPTLC1P2, GABBR1, MIR2113, EIF4EBP2P3, KMT2E |
| Bipolar[6] | 2024 | https://doi.org/10.1038/s41586-024-08468-9 | NEGR1, LINC01360, WDPCP, LOC100287010, CNTNAP5, TEX41, SATB1-AS1, TOPAZ1, NEK4, ITIH1, ITIH3, ITIH4, ITIH4-AS1, SUCLG2-AS1, TAFA1, FBXL17, PRR16, GPM6A, BTN3A2, BTN2A2, BTN3A1, HIVEP2, TMEM106B, PCLO, CYP7B1, LINC00051, TSNARE1, ZDHHC21, PTCH1, LOC100507346, SORCS3, SORCS3-AS1, ELP4, PAX6, PAX6-AS1, PAUPAR, MIR130A, YPEL4, CLP1, ZDHHC5, MED19, TMX2, TMX2-CTNND1, TMEM258, MIR611, FEN1,FADS1, MIR1908, FADS2, LOC101928847, NCAM1, IGSF9B, SOX5, SP7, SP1, AMHR2, LOC101929058, CIT, LINC00333, STK24, STK24-AS1, LRFN5, FCF1, YLPM1, IFT43, GPATCH2L, VAMP2, TMEM107, SNORD118, MIR4521, BORCS6, AURKB, LINC00324, CTC1, PFAS, ASXL3, RAB27B, MMP9, SLC12A5-AS1, SLC12A5, NCOA5, BRWD1, BRWD1-AS2, BRWD1-AS1, HMGN1, GET1, WRB-SH3BGR, SDC4P, SEC14L4, SEC14L6, GAL3ST1, PES1, RBX1, SNORD140+D5 |
| Bipolar 2[7] | 2021 | https://doi.org/10.1038/s41588-021-00857-4 | LINC01748, NUF2, LMAN2L, SCN2A, CERS6, PCGEM1, TRANK1, ITIH1, MDFIC2, CD47, KIAA1109, ADCY2, HOMER1, SSBP2, KDM3B, DOCK2, MHC, POU3F2, SYNE1, RPS6KA2, MAD1L1, THSD7A, SP4, MPP6, SRPK2, PLXNA4, MRPS33, miR124-1, MSRA, RP1-84O15.2, PLEC, ZCCHC7, TUBBP5, CACNB2, ANK3, ADO, ADD3, FADS2, FKBP2, PACS1, PC, SHANK2, CACNA1C, CUL4A, BCL11B, C15orf53, STARD9, HOMER2, ZNF592, FURIN, C16orf72, GRIN2A, RPL13, RTN4RL1, ERBB2, HDAC5, STK4, WFDC12, KCNB1, OSBPL2, SLC25A17, HTR6, DCLK3, HAPLN4, PACSIN2 |
| MDD[8] | 2025 | https://doi.org/10.1016/j.cell.2024.12.002 | RERE, RERE-AS1, ZMYM4, RP5-1180C18.1, RIMS3, ELAVL4, NRDC, NFIA, PDE4B, RPL31P12, LINC01725, PKN2-AS1, GAS5, COP1, RP1-35C21.2, CACNA1E, NAV1, IPO9-AS1, PROX1-AS1, USH2A, ESRRG, PSEN2, OPN3, SMYD3, RP11-254F7.2, NBAS, AC067959.1, AC068490.1, TRMT61B, LINC01320, NRXN1, ACTG1P22, FANCL, CNTNAP5, THSD7B, LRP1B, ARHGAP15, AC096558.1, TEX41, GALNT13, TANK, AC092684.1, SLC25A12, RAPGEF4, CIR1, ZNF804A, ERBB4, AC107218.3, NYAP2, RP11-314B1.2, GIGYF2, KIF1A, CNTN4, LMCD1-AS1, PLCL2, SATB1-AS1, ANO10, ZNF445, GPX1, CACNA2D3, PTPRG, MAGI1, SAMMSON, FOXP1, ROBO2, ZBTB20, LSAMP, RSRC1, KCNMB2, EEF1A1P23, OPA1, HTT, LDB2, KCNIP4, GABRB1, NFXL1, LINC01088, NAA11, LINC02267, EMCN, NDST3, BLTP1, MAML3, MARCHF1, GPM6A, TENM3, LINC01019, CDH18, CDH9, ARL15, SCAMP1, MEF2C-AS2, CTC-467M3.3, MEF2C, FAM172A, RP11-6N13.1, LINC02147, LINC02208, LINC02240, RP11-114J13.1, PCYOX1L, LINC01470, SGCD, LINC03000, TENM2, NPM1, IRF4, FARS2, LNC-LBCS, LINC00951, EYS, LIN28B, GOPC, NKAIN2, SAMD3, SAMD5, PCMT1, ESR1, ARID1B, PRKN, PACRG, MAD1L1, AC110781.3, AMZ1, SDK1, KDELR2, NXPH1, TMEM106B, ETV1, DGKB, TSPAN13, SP4, DNAH11, AVL9, ABCA13, AUTS2, CALN1, GTF2IRD1, MAGI2, PCLO, RP11-325F22.2, THAP5, DNAJB9, DOCK4, FOXP2, CTTNBP2, CADPS2, GRM8, CHCHD3, LRGUK, CNOT4, MKRN1, PTPRN2, CSMD1, SGCZ, PPP3CC, LINC01414, CYP7B1, ZFHX4, RP11-122C21.1, CCDC26, CARM1P1, PTPRD, LINC01235, RP11-536O18.1, RP11-132E11.2, ELAVL2, LINGO2, PAX5, CKS2, RP11-165J3.5, PTCH1, LINC01505, ASTN2, RABGAP1, NCS1, BRD3, EXD3, FBH1, SFMBT2, CAMK1D, CACNB2, CTNNA3, FUT11, LRMDA, ZMIZ1, SUFU, CNNM2, SORCS3, B4GALNT4, RP11-396O20.1, SOX6, LUZP2, LINC02755, ZDHHC5, FADS1, SHANK2, DLG2, GRM5, CNTN5, KIRREL3, RP11-168K9.2, AP002856.5, NTM, OPCML, B3GAT1-DT, PAUPAR, CACNA1C, ETV6, MGST1, LMO3, ST8SIA1, SOX5, CPNE8, LRRK2, GPR84-AS1, GPR84-AS1, RP11-644F5.16, TSFM, NAV3, RP11-328J6.1, UBE3B, ALDH2, NOS1, SPPL3, MPHOSPH9, ATP8A2, ALG5, DGKH, PCDH9, OBI1-AS1, NDFIP2, GPC5, GPC6, STK24, NPAS3, LRFN5-DT, CTD-2307P3.1, DCAF5, DLST, RP11-61O1.1, KLC1, RP11-73M18.2, RP11-73M18.7, SEMA6D, CTD-2050N2.1, MEGF11, LOXL1-AS1, LINGO1, NTRK3, FURIN, VPS33B-DT, AGBL1, RBFOX1, SNX29, SHISA9, RP11-1021N1.1, BMERB1, METTL9, CTB-31N19.3, ZNF423, FTO, GNAO1, PMFBP1, ZFHX3, CDH13, SLC7A5, METTL16, SHISA6, FLOT2, ASIC2, THRA, HOXB-AS3, BPTF, CCDC178, SETBP1, DCC, RAB27B, TCF4, ZCCHC2, RP11-154H12.3, TSHZ3-AS1, ZHX3, CD40, CDH22, ZMYND8, CSE1L, LINC01524, MIR646HG, CHRNA4, MYT1, CBR3, RSPH14, OSBP2, MIRLET7BHG, MAPK11 |
| PTSD[9] | 2024 | https://doi.org/10.1038/s41588-024-01707-9 | ZMYM4, PDE4B, CACNA1E, CNTNAP5, THSD7B, ARHGAP15, TANK, PLCL2, SNRK, FOXP1, ANAPC4, IPO11, EFNA5, FBXL17, GRIA1, SGCD, ASCC3, ESR1, SDK1, TMEM106B, SP4, PCLO, FOXP2, GRM8, TNKS, TSNARE1, FAM120A, OLFM1, C10orf11, SORCS3, MEN1, NCAM1, RP11-552I14.1, NOS1, PEBP1, SUDS3, LRFN5, GALNT16, TRAF3, LINGO1, C17orf58, BPTF, DCC, TCF4, LSM4, IGSF1, MTM1 |
| SUD[10] | 2023 | https://doi.org/10.1038/s44220-023-00034-y | PDE4B, GTF3C2, IFT172, GCKR, C2orf16, PLCL2, PRKAR2A, SLC25A20, ARIH2, P4HTM, WDR6, QRICH1, QARS, USP19, C3orf84, CCDC36, RP11-3B7.1, USP4, RHOA, NICN1, DAG1, ADD1, NOP14, BANK1, PPP6C, SOX6, BDNF, MTCH2, ZDHHC5, TMX2, TMX2-CTNND1, C11orf31, RP11-691N7.6, CTNND1, ANKK1, DRD2, HS6ST3, ARID4A, PPP1R13B, SEMA6D, FTO, C20orf112, ADH1B, FUT2, C4ORF54 |

**Table S7.**
*Genes underpinning PRS disorder scores, extracted from relevant GWAS analyses*
Relevant GWAS analysis are referenced and presented along with lists of genes shared in their original publications, applied in gene ontology analysis.
